# Estimating epidemiologic dynamics from cross-sectional viral load distributions

**DOI:** 10.1101/2020.10.08.20204222

**Authors:** James A. Hay, Lee Kennedy-Shaffer, Sanjat Kanjilal, Niall J. Lennon, Stacey B. Gabriel, Marc Lipsitch, Michael J. Mina

**Affiliations:** Center for Communicable Disease Dynamics, Harvard T H Chan School of Public Health, Boston, MA.; Department of Epidemiology, Harvard T H Chan School of Public Health, Boston, MA.; Department of Immunology and Infectious Diseases, Harvard T H Chan School of Public Health, Boston, MA.; Department of Mathematics and Statistics, Vassar College, Poughkeepsie, NY.; Department of Population Medicine, Harvard Pilgrim Health Care Institute, Boston, MA.; Department of Infectious Diseases, Brigham and Women’s Hospital, Boston, MA.; Broad Institute of MIT and Harvard, Cambridge, MA.; Department of Pathology, Brigham and Women’s Hospital, Boston, MA.

## Abstract

Estimating an epidemic’s trajectory is crucial for developing public health responses to infectious diseases, but incidence data used for such estimation are confounded by variable testing practices. We show instead that the population distribution of viral loads observed under random or symptom-based surveillance, in the form of cycle threshold (Ct) values, changes during an epidemic and that Ct values from even limited numbers of random samples can provide improved estimates of an epidemic’s trajectory. Combining multiple such samples and the fraction positive improves the precision and robustness of such estimation. We apply our methods to Ct values from surveillance conducted during the SARS-CoV-2 pandemic in a variety of settings and demonstrate new approaches for real-time estimates of epidemic trajectories for outbreak management and response.

## Main Text

Real-time tracking of infection incidence during an epidemic is fundamental for public health planning and intervention (*1, 2*). In the severe acute respiratory syndrome coronavirus-2 (SARS-CoV-2) pandemic, key epidemiological parameters such as the time-varying effective reproductive number, *R*_*t*_, have typically been estimated using the time-series of observed case counts, percent of positive tests, or deaths, usually based on reverse-transcription quantitative polymerase chain reaction (RT-qPCR) testing. However, reporting delays (*3*), limited testing capacities, and changes in test availability over time all impact the ability of routine testing to reliably and promptly detect underlying changes in infection incidence (*4, 5*). In particular, whether changes in case counts at different times reflect epidemic dynamics or simply changes in testing have been major topics of debate with important economic, health and political ramifications. Here, we describe a new method to overcome these biases and obtain accurate estimates of the epidemic trajectory, one that does not require repeat measurements and uses routinely generated but currently discarded quantitative data from RT-qPCR testing from single or successive cross-sectional samples.

RT-qPCR tests provide quantitative results in the form of cycle threshold (Ct) values, which are inversely correlated with log_10_ viral loads, but they are often reported only as binary “positives” or “negatives” (*6, 7*). It is common when testing for other infectious diseases to use this quantification of sample viral load, for example, to identify individuals with higher clinical severity or transmissibility (*8–11*). For SARS-CoV-2, Ct values may be useful in clinical determinations about the need for isolation and quarantine (*7, 12*), identifying the phase of an individual’s infection (*13, 14*) and predicting disease severity (*14, 15*). However, individual-level decision making based on Ct values has not yet become a widespread reality due to the variability in measurements across testing platforms and samples, and limited data to understand SARS-CoV-2 viral kinetics in asymptomatic and presymptomatic infections. These concerns do not necessarily hold at the population level: whereas a single high Ct value may not necessarily guarantee a low viral load in one sample, high Ct values in many samples will indicate a population with predominantly low viral loads. Indeed, the population-level distribution of Ct values does appear to change over time. For example, a systematic incline in the distribution of quantified Ct values has been noted alongside epidemic decline (*12, 14, 16*).

We demonstrate that population-level changes in the distribution of observed Ct values can arise as an epidemiological phenomenon, and propose methods to use these quantitative values to estimate epidemic trajectories from one or more cross-sectional samples.

### Relationship Between Observed Ct Values and Epidemic Dynamics

First, we show that the interaction of within-host viral kinetics and epidemic dynamics can drive changes in the distribution of Ct values over time without a change in the underlying pathogen kinetics. To demonstrate the epidemiological link between transmission rate and measured viral loads or Ct values, we first simulated infections arising under a deterministic susceptible-exposed-infectious-recovered (SEIR) model (Fig. 1A, *Materials and Methods: Simulated Epidemic Transmission Models)*. Parameters used are in Table S1. At selected testing days during the outbreak, simulated Ct values are observed from a random sample of the population using the Ct distribution model described in *Materials and Methods: Ct Value Model* and shown in Figs. S1 and S2. By drawing simulated samples for testing from the population at specific time points, these simulations recreate realistic cross-sectional distributions of detectable viral loads across the course of an epidemic. Throughout, we assume each individual is infected at most once, ignoring re-infections as these appear to be a negligible portion of infections in the epidemic so far (*17*).

**Fig. 1.**
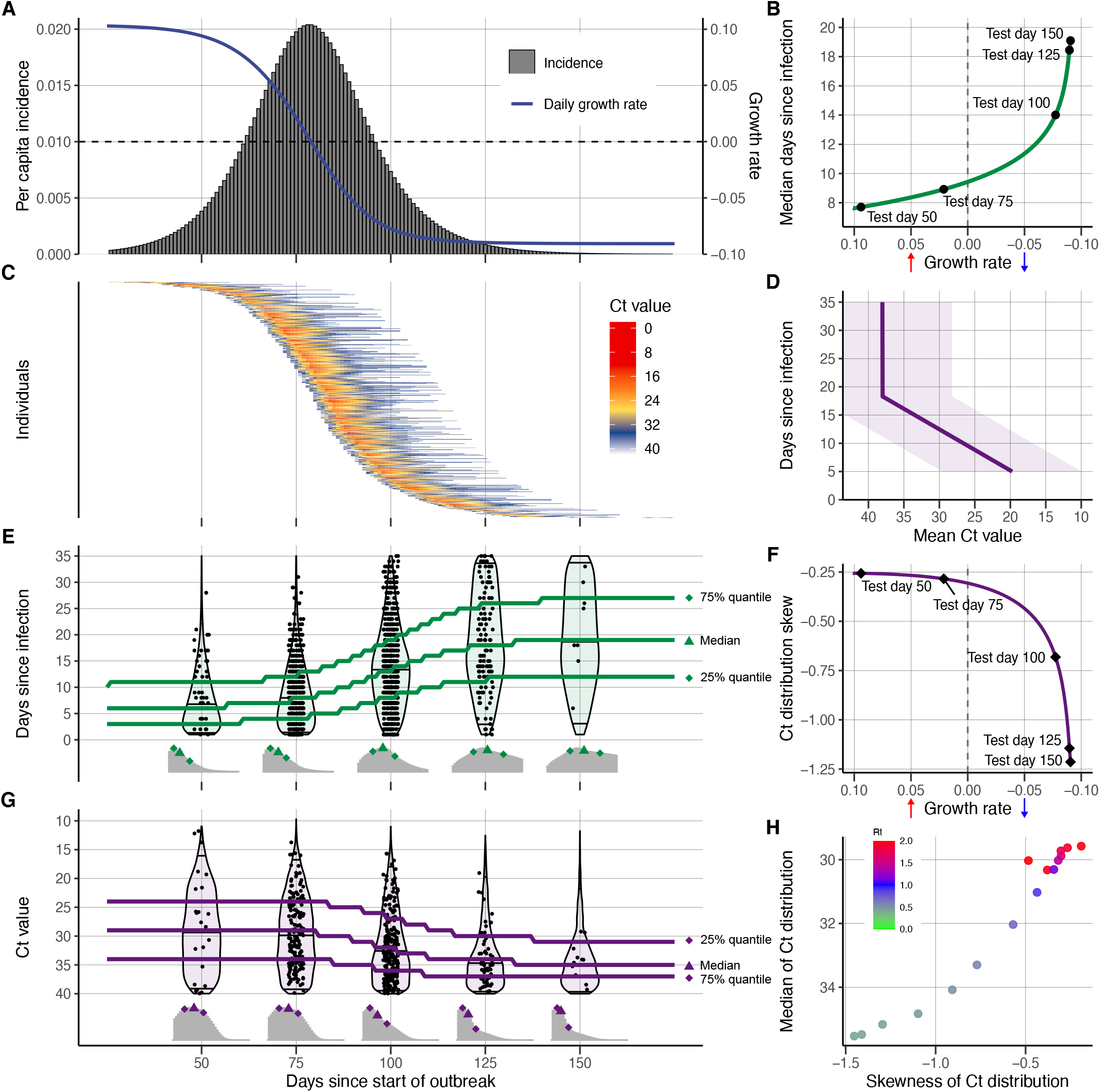
The cycle threshold (Ct) value distribution reflects epidemiological dynamics over the course of an outbreak. (**A**) Per capita daily incidence (histogram) and daily growth rate (blue line) of new infections in a simulated epidemic using a susceptible-exposed-infectious-recovered (SEIR) model. (**B**) Median days since infection vs. daily growth rate of new infections by epidemic day. Labeled points here and in (**E–G**) show five time points in the simulated epidemic. (**C**) Observed Ct value by day for 500 randomly sampled infected individuals. (**D**) Viral kinetics model (increasing Ct value following peak and subsequent plateau near the limit of detection), demonstrating the time course of Ct values (x-axis, line shows mean and ribbon shows 95% quantile range) against days since infection (y-axis). Note that the y-axis is arranged to align with (**E**). (**E**) Distribution of days since infection (violin plots and histograms) for randomly selected individuals over the course of the epidemic. Median and first and third quartiles are shown as green lines and points. (**F**) Skewness of observed Ct value distribution vs. daily growth rate of new infections by epidemic day. (**G**) Distribution of observed Ct values (violin plots and histograms) among sampled infected individuals by epidemic day. Median and first and third quartile are shown as purple lines and points. (**H**) Time-varying effective reproductive number, *R*_*t*_, derived from the SEIR simulation, plotted against median and skewness of observed Ct value distribution.

Early in the epidemic, infection incidence grows rapidly and the typical infection is thus recent; as the epidemic wanes, however, the average time since exposure increases as the rate of new infections decreases (Fig. 1B,E) (*18*); this is analogous to the average age being lower in a growing vs. declining population (*19*). Infections are often unobserved events, but we can rely on an observable quantity, such as viral load, as a proxy for the time since infection. Since Ct values change over time within infected hosts (Fig. 1C), random sampling of individuals during epidemic growth is more likely to measure individuals who were recently infected and therefore in the acute phase of their infection with higher quantities of viral RNA. Conversely, sampling infected individuals during epidemic decline is more likely to capture individuals in the convalescent phase, typically sampling lower quantities of viral RNA (Fig. 1D). The distribution of observed Ct values therefore changes over time, as measured by the median, quartiles, and skewness (Fig. 1G). While estimates for an individual’s time since infection based on a single Ct value will be highly uncertain, the population-level distribution of observed Ct values will vary with the growth rate, and therefore *R*_*t*_, of new infections (Fig. 1F,H). Similar principles have been applied to serologic data to infer unobserved individual-level infection events (*16, 20–22*) and population-level parameters of infectious disease spread (*20, 23–27*).

This phenomenon is also present, though less pronounced, among viral loads measured under symptom-based surveillance (Fig. S3). One might imagine that the typical time since infection would not depend on the epidemic trajectory in individuals systematically sampled soon after symptom onset. However, the distribution of delays between infection date and test date is a convolution of the infection incidence curve and the confirmation delay distribution (time from infection to testing of symptomatic infections) (*28*). Individuals tested due to recent symptom onset are more likely to have been recently infected with a short incubation period during epidemic growth than during epidemic decline, where more onsets are from older infections with longer incubation periods. The time-since-infection distribution of individuals tested based on symptom onset, and therefore their measured viral loads, is influenced by the stage of the epidemic.

By modeling the variation in observed Ct values arising from individual-level viral growth/clearance kinetics and sampling errors, the distribution of observed Ct values becomes an estimable function of the times since infection, and the expected median and skewness of Ct values at a given point in time are then predictable from the growth rate. This function can then be used to estimate the epidemic growth rate conditional on a set of observed Ct values. The relationship between observed Ct value and epidemic growth rate holds for any testing approach, though calibration is needed to define the precise mapping (i.e., using a different RT-qPCR instrument, a different Ct value threshold, or in a different lab; see Fig. S4).

### Inferring Epidemic Trajectory Using a Single Cross-Section

From these relationships, we derived a method to formally infer the epidemic growth rate given a single cross-section of RT-qPCR test results. The method combines two models: (1) the likelihood of observing a Ct value or negative result conditional on having been infected on a given day; and (2) the likelihood of being infected on a given day prior to the sample date. For (1), we used a Bayesian model and defined priors for the mode and range of Ct values following infection based on the existing literature (*Materials and Methods: Ct Value Model* and *Single Cross-Section Model*). For (2), we initially developed two models to describe the probability of infection over time: (a) constant exponential growth of infection incidence; or (b) infections arising under an SEIR model. Both models provide estimates for the epidemic growth rate, but make different assumptions regarding the possible shape of the outbreak trajectory: the exponential growth model assumes a constant growth rate, whereas the SEIR model assumes that the growth rate changes daily depending on the remaining number of susceptible individuals.

We first investigated how the distribution of Ct values and prevalence of PCR positivity changed over time in four well-observed Massachusetts long-term care facilities that underwent SARS-CoV-2 outbreaks in March and April 2020 (*29*). These facilities were relatively closed after outbreaks began, so we model the outbreak within each facility using an extended SEIR (SEEIRR) model, with additional exposed and recovered compartments to account for the duration of PCR positivity (*Materials and Methods: Simulated Epidemic Transmission Models*). In each facility, we have the results of near-universal PCR testing, including both residents and staff, from three time points after the outbreak began, including the number of positive samples, the Ct values of positive samples, and the number of negative samples (*Materials and Methods: Nursing Home Data*). Fig. 2 shows results for one of these facilities, while Fig. S5 shows results for the other three.

**Fig. 2.**
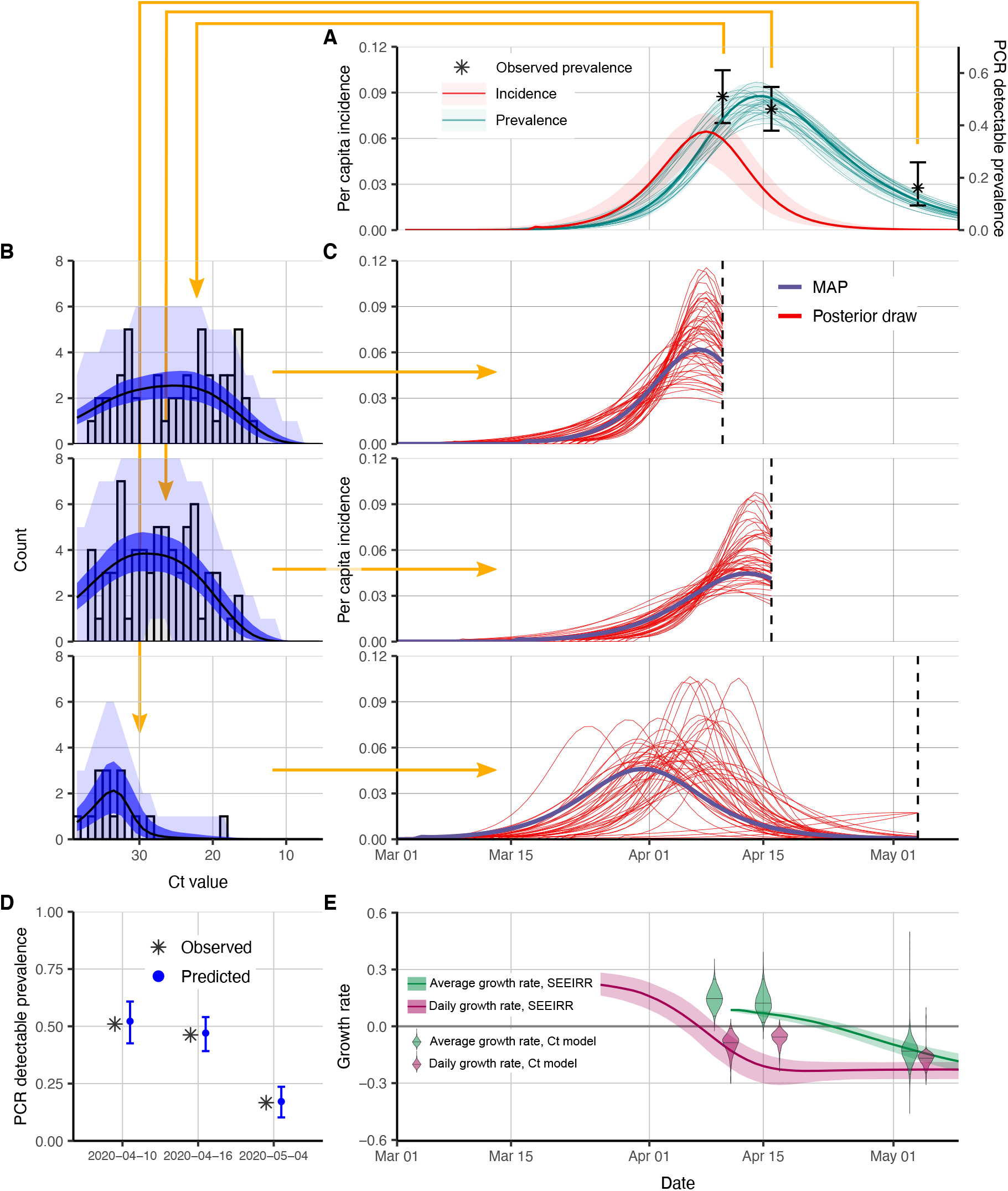
Single cross-sectional distributions of observed cycle threshold (Ct) values can be used to reconstruct epidemic trajectories in a Massachusetts nursing home. (**A**) Estimated prevalence (faint teal lines show posterior samples, solid teal line shows posterior median, teal ribbon shows 95% CrI) and incidence (red line shows posterior median, red ribbon shows 95% CrI) from the standard compartmental (SEEIRR) model fit to point prevalence at three sampling times (error bars show 95% binomial confidence intervals). (**B**) Model-predicted Ct distributions (blue) fitted to the observed Ct values (grey bars) from each of three cross-sectional samples. Shown are the posterior median (black line) and 95% CrI for the expected Ct distribution (dark blue ribbon), and 95% prediction intervals based on simulated observations (light blue ribbon). Note that prediction intervals are much wider than credible intervals, as they result from simulating observations with a small sample size. (**C**) Each panel shows results from fitting the Ct-based SEIR model separately to three cross-sections of virologic data. Shown are random posterior samples (red lines) and the maximum posterior probability trajectory (purple line) for the incidence curve. (**D**) Ct model-predicted median (blue point) and 95% CrI (blue error bars) for the proportion of samples testing positive compared to the observed proportion tested positive (grey cross). (**E)** 35-day (green) and 1-day (magenta) average growth rates from the Ct model estimates in part (**C**) at three time points (violin plots) compared to growth rate estimates from the SEEIRR model in part (**A**) (lines and shaded ribbons).

In Fig. 2A, we fit the SEEIRR compartmental model to the three observed point prevalence values from the facility as a benchmark. The distribution of observed Ct values at each time point (Fig. 2B) shifts higher and becomes more left-skewed at later time points. We then fit the exponential growth and simple SEIR models using the Ct likelihood to each individual cross-section to get posterior distributions for the epidemic trajectory up to that time point (Fig. 2C). Note that these fits do not use any longitudinal data; each is fit to the positive and negative Ct values from only one time point. To assess the fit, we compare the predicted Ct distribution and point prevalence from each fit to the data (Fig. 2B,D) and compare the growth rates from these fits to those derived from the fits to the point prevalences. Posterior distributions of all Ct value model parameters are shown in Fig. S6.

While both sets of results are fitted models and so neither can be considered the truth, we find that the Ct method fit to one cross-section of data provides a similar posterior median trajectory to the compartmental model fit to three point prevalences. In particular, the Ct-based models appear to accurately discern whether the samples were taken soon or long after peak infection incidence. Both methods were in agreement over the direction of the past average and recent daily growth rates (i.e., whether the epidemic is currently growing or declining, and whether the growth rate has dropped relative to the historic average). The average growth rate estimates were very similar at most time points, though the daily growth rate appeared to decline earlier in the compartmental model. Overall, these results demonstrate that a single cross-section of Ct values can provide similar information to point prevalence estimates from three distinct sampling rounds.

To ensure that our method provides accurate estimates of the epidemic trajectory, we performed extensive simulation-recovery experiments using a synthetic nursing home population undergoing a stochastic SEIR epidemic. We assess performance using various models, including a version that uses only positive Ct values, and varying parameters of the simulation; details are in *Materials and Methods: Simulated Nursing Home Outbreaks* and results in Figs. S7–S9.

### Inferring Epidemic Trajectory Using Multiple Cross-Sections

Next, we extended our method to combine data from multiple cross-sections, allowing us to more reliably estimate the epidemic trajectory (*Materials and Methods: Multiple Cross-Sections Model* and *Markov Chain Monte Carlo Framework*). In many settings, the epidemic trajectory is monitored using reported case counts, the definition of which can change during the epidemic (*30*). Limiting reported cases to positive test results, the number of new positives among the tests conducted each day can be used to calculate *R*_*t*_ (*3*). However, these data represent the growth rate of positive tests and not the incidence of infection, requiring adjustments to account for changes in testing capacity, the delay between infection and test report date, and the conversion from prevalence to incidence. When, instead, Ct values from surveillance sampling is available, our methods can overcome these limitations by providing a direct mapping between the distribution of Ct values and infection incidence. Crucially, the Ct-based methods are agnostic to changing testing rates, providing unbiased growth rate estimates where case count-based methods exhibit bias (*5*).

To demonstrate the performance of these methods, we use them to recover parameters from SEIR-based simulations under a variety of testing schemes (*Materials and Methods: Simulated Testing Schemes*). We compare the performance of *R*_*t*_ estimation using reported case counts via the R package *EpiNow2* (*28, 31*), where reporting depends on testing capacity and the symptom status of infected individuals, to the performance of our methods when one, two, or three surveillance samples are available with observed Ct values, with a total of about 0.3% of the population sampled (3000 tests spread among the samples).

Figure 3 plots the posterior median *R*_*t*_ from each of the 100 simulations of each method when the epidemic is growing (day 60) and declining (day 88). Except when only one sample is used, the Ct-based methods fitting to an SEIR model exhibit minimal bias, even when testing capacity changes. Methods based on reported case counts, on the other hand, exhibit noticeable upward bias when testing rates increase over the period observed and substantial downward bias when testing rates decrease. The Ct-based methods do exhibit higher variability, however. This is captured by the Bayesian inference model, as all of the Ct-based methods achieve at least nominal coverage of the 95% credible intervals among these 100 simulations. The methods based on reported case counts have coverage below 70% when testing is falling at either time point and when cases are rising while the epidemic is declining.

**Fig. 3.**
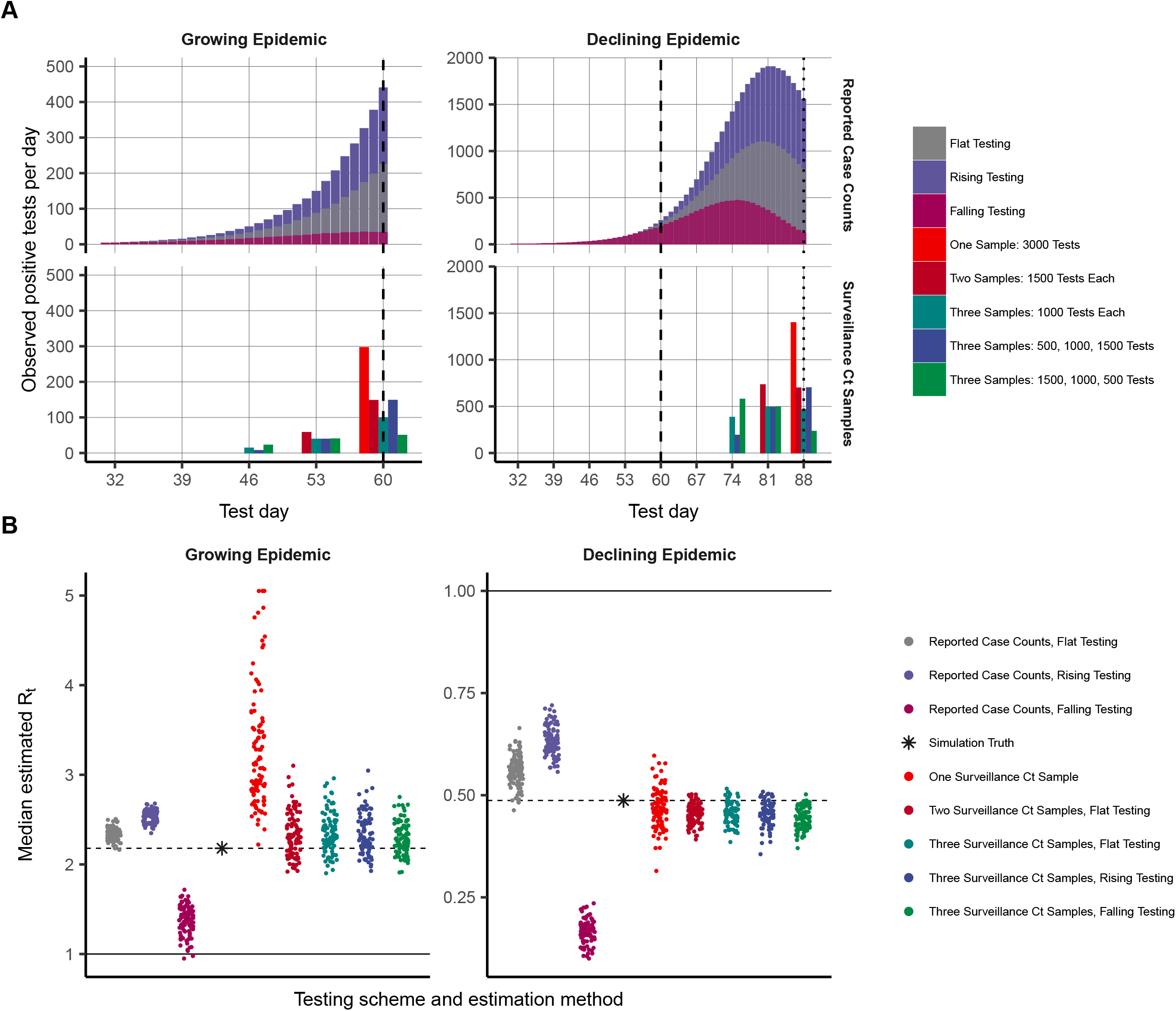
Inferring epidemic trajectory from cross-sectional surveillance samples with observed cycle threshold (Ct) values yields nearly unbiased estimates of the time-varying effective reproductive number, *R*_*t*_, whereas changing testing rates lead to biased estimation using reported case counts. (**A**) Number of positive tests per day by sampling time in epidemic and testing scheme for reported case counts (top row) and surveillance Ct sampling (bottom row), from a simulated susceptible-exposed-infectious-recovered (SEIR) epidemic. Observation times are shown by vertical lines. (**B**) *R*_*t*_ estimates from 100 simulations for each epidemic sampling time, testing scheme, and estimation method. Each point is the posterior median from a single simulation. *R*_*t*_ estimates for reported case counts use *EpiNow2* estimation and for surveillance Ct samples use the Ct-based likelihood for one or multiple cross-sections fitted to an SEIR model. True model-based *R*_*t*_ on the sampling day is indicated by the black star and dashed horizontal line, while an *R*_*t*_ of 1, indicating a flat outbreak, is indicated by the solid horizontal line.

### Reconstructing Complex Incidence Curves Using Ct Values

Simple epidemic models are useful to understand recent incidence trends when data are sparse or in relatively closed populations where the epidemic start time is approximately known (*Materials and Methods: Epidemic Seed Time Priors*). In reality, however, the epidemic usually follows a more complex trajectory which is difficult to model parametrically. For example, the SEIR model does not account for the implementation/relaxation of non-pharmaceutical interventions unless explicitly specified in the model. For a more flexible approach to estimating the epidemic trajectory from multiple cross-sections, we developed a third model for infection incidence, using a Gaussian Process (GP) prior for the underlying daily probabilities of infection (*32*). The GP method provides estimated daily infection probabilities without making strong assumptions about the epidemic trajectory, assuming only that infection probabilities on contemporaneous days are correlated, with decreasing correlation at increasing temporal distances (*Materials and Methods: Gaussian Process Model*). Movie S1 demonstrates how estimates of the full epidemic trajectory can be sequentially updated using this model as new samples become available over time.

With the objective of reconstructing the entire incidence curve using routinely collected RT-qPCR data, we used anonymized, Ct values from positive samples measured from nearly all hospital admissions into Brigham & Women’s Hospital (BWH) in Boston, MA, between April 3 and November 10, 2020 (*Materials and Methods: Brigham & Women’s Hospital Data*). We aligned these with estimates for *R*_*t*_ based on case counts in Massachusetts (Fig. 4A–C). The median and skewness of the detectable Ct distribution was correlated with *R*_*t*_ (Fig. 4B), in line with our theoretical predictions. Tests taken prior to April 3 were restricted to symptomatic patients only, while those after April 15 represented near-universal testing of all hospital admissions and non-admitted ER patients. The median Ct value rose (corresponding to a decline in median viral load) and skewness of the Ct distribution fell in the late spring and early summer, as shelter-in-place orders and other non-pharmaceutical interventions were rolled out (Fig. 4C), but the median declined and skewness rose in late summer and early fall as these measures were relaxed, coinciding with an increase in observed case counts for the state (Fig. 4A).

**Fig. 4.**
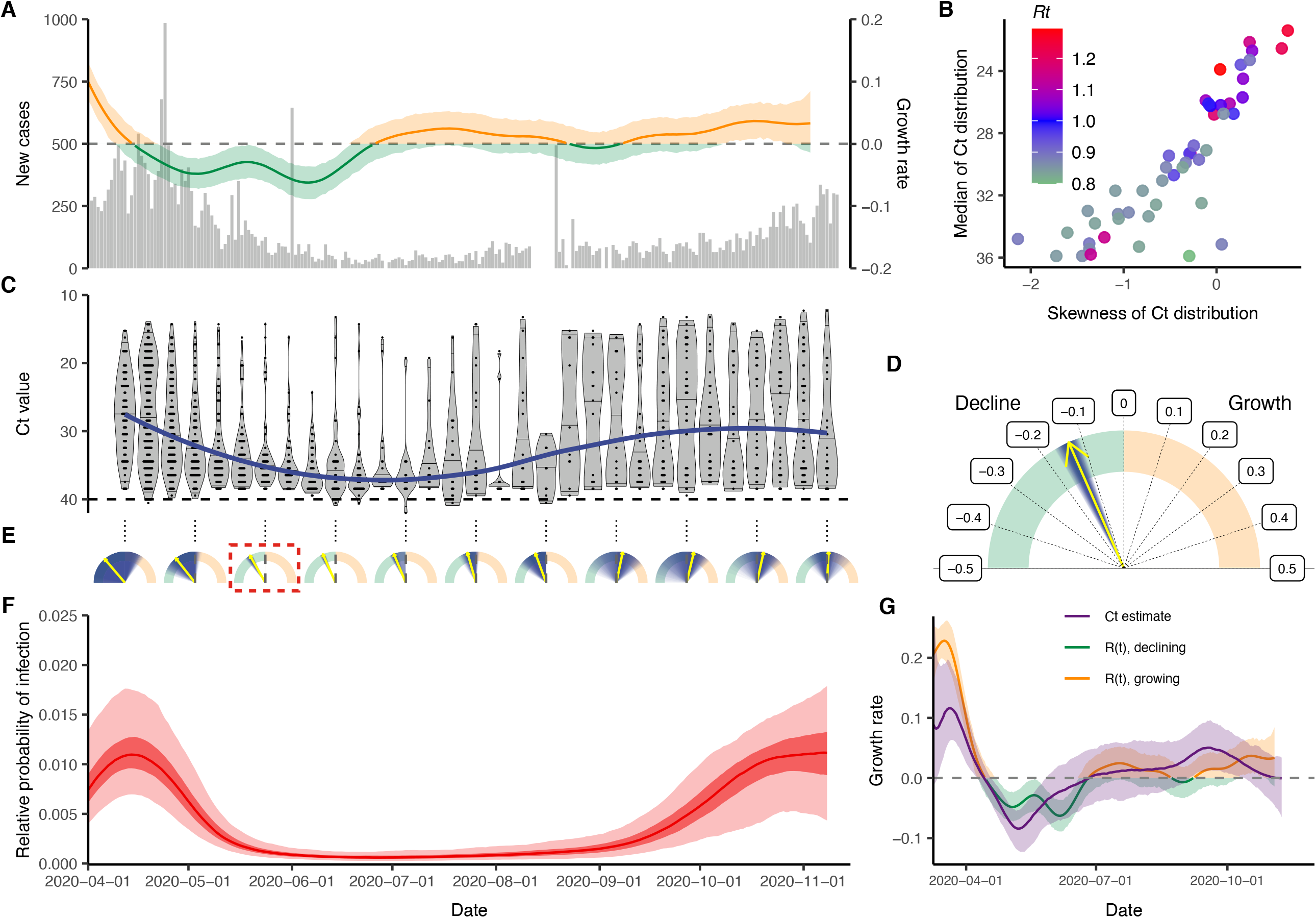
Single cross-sectional distributions of observed cycle threshold (Ct) values can estimate growth rate and multiple cross-sectional distributions can estimate the complex statewide epidemic trajectory from hospital-based surveillance at Brigham & Women’s Hospital in Massachusetts. (**A**) Daily confirmed new cases in Massachusetts (gray bars) and estimated time-varying effective reproductive number, *R*_*t*_. (**B**) Estimated *R*_*t*_ from the case counts vs. median and skewness of observed Ct value distribution by weekly sampling times. (**C**) Distribution (violin plots and points) and smoothed median (blue line) of observed Ct values by sampling week. (**D**) Posterior median (yellow arrow) and distribution (blue shaded area) of estimated daily growth rate of incident infections from a susceptible-exposed-infectious-recovered (SEIR) model fit to a single cross-section of observed Ct value data from the week commencing 2020-05-24. Shading density is proportional to posterior density. (**E**) Posterior medians (yellow arrow) and full distributions (blue shaded area) of estimated daily growth rate of incident infections from SEIR models each fit to a single cross-section by sampling week used. Red box denotes the panel from (**D**). (**F**) Posterior distribution of relative probability of infection by date from a Gaussian Process (GP) model fit to all observed Ct values (ribbons show 95% and 50% credible intervals, line shows posterior median). (**G**) Comparison of estimated daily growth rate of incident infections from GP model (blue line and shaded ribbons show posterior median and 95% CrI) to that from *R*_*t*_ estimation using observed case counts (red and green line and shaded ribbons show posterior median and 95% CrI) by date. Note that the x-axis is truncated at 2020-04-01, but estimates stretch back to 2020-03-01 (Fig. S13).

Using the observed Ct values we estimated the daily growth rate of infections using the SEIR model on single cross-sections (Fig. 4D,E, Fig. S10, Fig. S11) and the daily relative probability of infection over time using the GP model (Fig. 4F, Fig. S12). Similar temporal trends were inferred under both models, and the GP model provided growth rate estimates that followed those estimated using observed case counts (Fig. 4G). While these data are not strictly a random sample of the community, and the observed case counts do not necessarily provide a ground truth for the *R*_*t*_ value, this demonstrates the ability of this method to re-create epidemic trajectories and estimate growth or decline of cases using only positive Ct values collected through routine testing. Interestingly, our estimated epidemic trajectory using only routinely generated Ct values from a single hospital was remarkably similar to changes in viral loads obtained from wastewater data (Fig. S13) (*33*).

## Discussion

The usefulness of Ct values for public health decision making is currently the subject of much discussion and debate. One unexplained observation which has been consistently observed in many locations is that the distribution of observed Ct values has varied over the course of the current SARS-CoV-2 pandemic, which has led to questions over whether the fitness of the virus has changed (*12, 14, 16*). Our results demonstrate instead that this can be explained as a purely epidemiologic phenomenon, without any change in individual-level viral dynamics or testing practices. We find that properties of the population-level Ct distribution strongly correlate with estimates for the effective reproductive number or growth rate in real-world settings, in line with our theoretical predictions.

Using quantitative diagnostic test results from multiple different tests conducted in a single cross-sectional survey, Rydevik et al. (*18*) demonstrated that epidemic trends could be inferred from virologic data. The methods we describe here use the phenomenon observed in the present pandemic and the relationship between incidence rate, time since infection, and virologic test results to estimate a community’s position in the epidemic curve, under various models of epidemic trajectories, based on data from one or more cross-sectional surveys using a single virologic test. Despite the challenges of sampling variability, individual-level differences in viral kinetics, and limitations in comparing results from different laboratories or instruments, our results demonstrate that RT-qPCR Ct values, with all of their quantitative variability for an individual, can be highly informative of population-level dynamics. This information is lost when measurements are reduced to binary classifications.

Our results demonstrate that this method can be used to estimate epidemic growth rates based on data collected at a single time point, and independent of assumptions about the intensity of testing. Comparisons of simulated Ct values and observed Ct values with growth rates and *R*_*t*_ estimates validate this general approach. Results should be interpreted with caution in cases where the observed Ct values are not from a population census or a largely random sample, or when there are very few samples with detectable viral load. When testing is based primarily on the presence of symptoms or follow-up of contacts of infected individuals, people may be more likely to be sampled at specific times since infection and thus the distribution of observed Cts would not be representative of the population as a whole. This method may be most useful in settings where representative surveillance samples can be obtained independent of COVID-19 symptoms, such as the REACT study (*34*). These methods allow municipalities to evaluate and monitor, in real-time, the role of various epidemic mitigation interventions, for example by conducting even a single or a small number of random virologic testing samples as part of surveillance.

These results are sensitive to the true distribution of observed viral loads each day after infection. Different swab types, sample types, instruments, or Ct thresholds may alter the variability in the Ct distribution (*15, 16, 35, 36*), leading to different relationships between the specific Ct distribution and the epidemic trajectory. Setting-specific calibrations, for example based on a reference range of Ct values, will be useful to ensure accuracy. Here, we generated a viral kinetics model based on observed properties of measured viral loads (proportion detectable over time following symptom onset, distribution of Ct values from true specimens), and used these results to inform priors on key parameters when estimating growth rates. The growth rate estimates can therefore be improved by choosing more precise, accurate priors relevant to the observations used during model fitting. In cases where results come from multiple testing platforms, the model should either be adjusted to account for this by specifying a different distribution for each platform based on its properties or, if possible, the Ct values should be transformed to a common scale such as log viral copies. Results could also be improved if individual-level features that may affect viral load, such as symptom status, age, and antiviral treatment, are available with the data and incorporated into the Ct value model (*14–16, 37, 38*). A similar approach may also be possible using serologic surveys, as an extension of work that has related time since infection to antibody titers for other infectious diseases (*26, 27*). If multiple types of tests (e.g., antigen and PCR) are conducted at the same time, combining information can substantially reduce uncertainty in these estimates as well (*18*). If variant strains are associated with different viral load kinetics and become common (*39*), this should be incorporated into the model as well.

This method has a number of limitations. While the Bayesian framework incorporates the uncertainty in viral load distributions into inference on the growth rate, parametric assumptions and reasonably strong priors on these distributions aid in identifiability. If these parametric assumptions are violated, inference may not be reliable. In addition, the methods described here and the relationship between incidence and skewness of Ct distributions become unreliable when there are very few positive cases, so results should be interpreted with caution and sample sizes increased in periods with low incidence. In some cases with one or a small number of cross-sections, the observed Ct distribution could plausibly result from all individuals very early in their infection at the start of fast epidemic growth, all during the recovery phase of their infection during epidemic decline, or a mixture of both (Fig. 4E, Fig. S11). We therefore used a parallel tempering MCMC algorithm which is able to accurately estimate these multimodal posterior distributions. Interpretation of the estimated median growth rate and credible intervals should be done with proper epidemiological context: estimated growth rates that are grossly incompatible with other data can be safely excluded.

This method may also overstate uncertainty in the viral load distributions if results from different machines or protocols are used to inform the prior. A more precise understanding of the viral load kinetics, and modeling those kinetics in a way that accounts for the epidemiologic and technical setting of the measurements, will help improve this approach and determine whether Ct distribution parameters from different settings are comparable. Because of this, quantitative measures from RT-qPCR should be reported regularly for SARS-CoV-2 cases and early assessment of pathogen load kinetics should be a priority for future emerging infections. The use of a control procedure in the measurements, like using the ratio of detected viral RNA to detected human RNA, could also improve the reliability and comparability of Ct measures.

The Ct value is a measurement with magnitude, which provides information on underlying viral dynamics. Although there are challenges to relying on single Ct values for individual-level decision making, the aggregation of many such measurements from a population contains substantial information. These results demonstrate how population-level distributions of Ct values can provide information on important epidemiologic questions of interest, even from a single cross-sectional survey. Better epidemic planning and more targeted epidemiological measures can then be implemented based on this survey, or use of Ct values can be combined with repeated sampling to maximize the use of available evidence.

## Data Availability

All code to perform the analyses and generate the figures presented in this article is available at https://github.com/jameshay218/virosolver_paper and https://github.com/jameshay218/virosolver. Simulated data and real data used in the analyses are also available at https://github.com/jameshay218/virosolver_paper. For the model fitting, code for the Markov chain Monte Carlo framework is available at https://github.com/jameshay218/lazymcmc and https://github.com/jameshay218/lazymcmc/tree/parallel_tempering. The authors used code developed by Abbott et al. to estimate Rt from reported case counts; this is available at https://github.com/epiforecasts/EpiNow2.

## Acknowledgments

We thank Steven Riley for helpful discussions.

## Funding

U.S. National Institutes of Health Director’s Early Independence Award DP5-OD028145 (MJM, JAH)

Morris-Singer Fund (LKS, ML)

U.S. Centers for Disease Control and Prevention Award U01IP001121 (LKS, ML)

U.S. National Institute of General Medical Sciences award U54GM088558 (ML, JAH, LKS)

## Author contributions

Conceptualization: JAH, LKS, ML, MJM

Methodology: JAH, LKS, ML, MJM Visualization: JAH, LKS, ML, MJM

Investigation: JAH, LKS, SK, ML, MJM

Resources: SK, NJL, SBG, MJM

Data curation: JAH, SK, NJL, SBG, MJM

Software: JAH, LKS, SK, NJL, SBG

Funding acquisition: ML, MJM

Supervision: ML, MJM

Writing – original draft: JAH, LKS, SK, ML, MJM

Writing – review & editing: JAH, LKS, SK, NJL, SBG, ML, MJM

## Competing interests

ML discloses honoraria/consulting from Merck, Affinivax, Sanofi-Pasteur, and Antigen Discovery; research funding (institutional) from Pfizer, and unpaid scientific advice to Janssen, Astra-Zeneca, and Covaxx (United Biomedical). MJM is a medical advisor for Detect. All other authors declare no competing interests.

## Data and materials availability

All code to perform the analyses and generate the figures presented in this article is available at https://github.com/jameshay218/virosolver_paper and https://github.com/jameshay218/virosolver. Simulated data and real data used in the analyses are also available at https://github.com/jameshay218/virosolver_paper. For the model fitting, code for the Markov chain Monte Carlo framework is available at https://github.com/jameshay218/lazymcmc and https://github.com/jameshay218/lazymcmc/tree/parallel_tempering. The authors used code developed by Abbott et al. to estimate *R*_*t*_ from reported case counts; this is available at https://github.com/epiforecasts/EpiNow2.

---

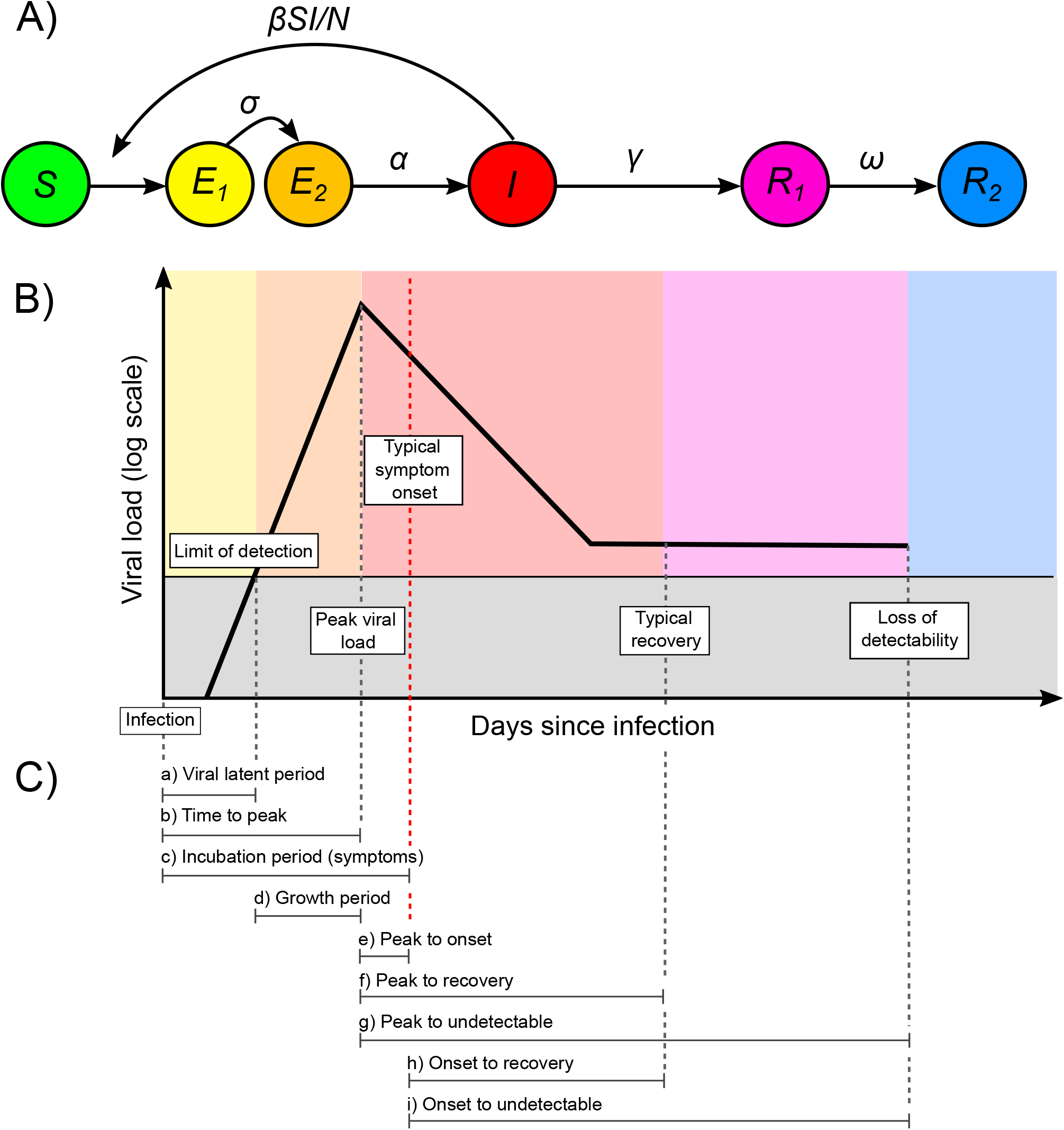

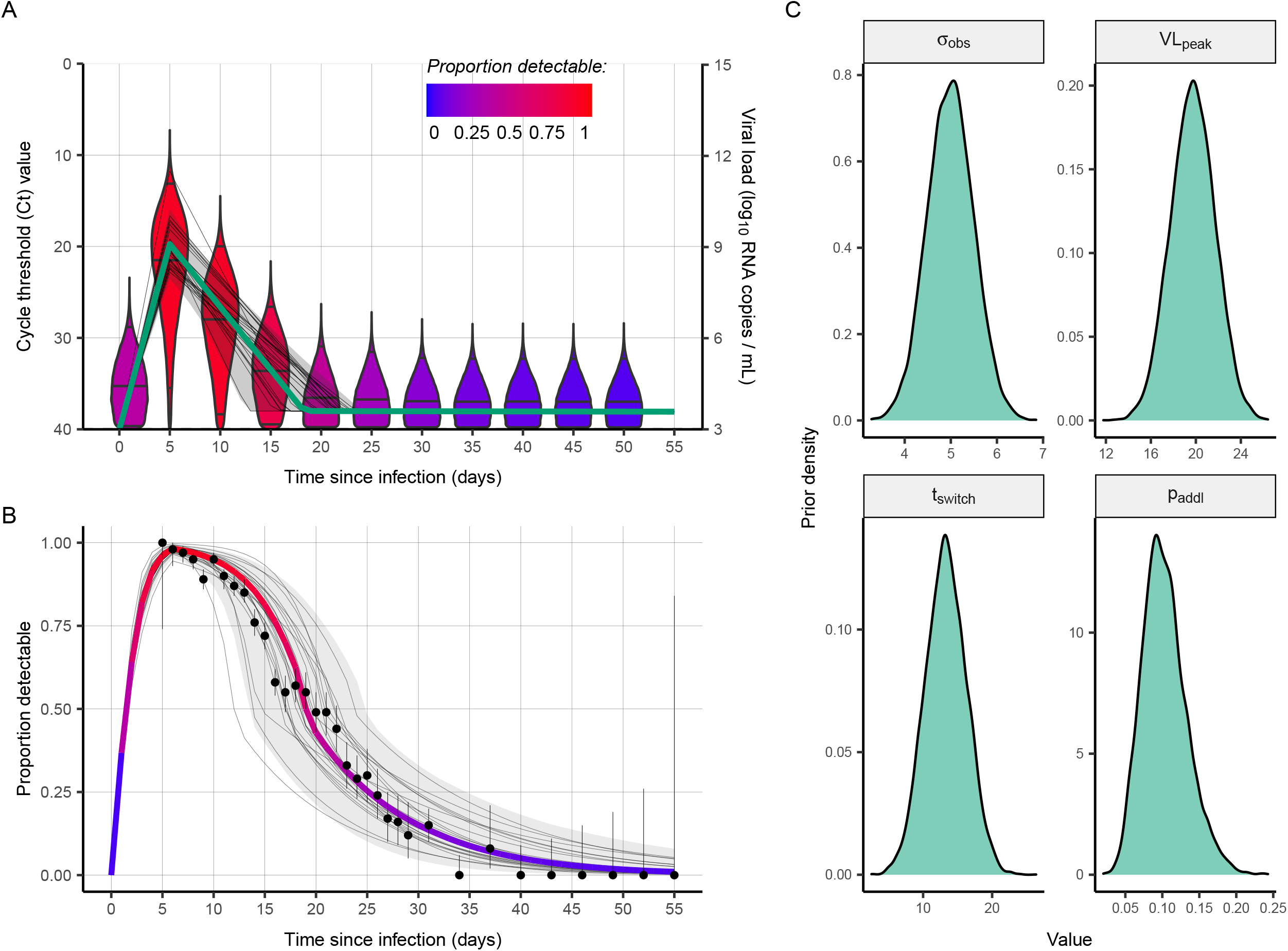

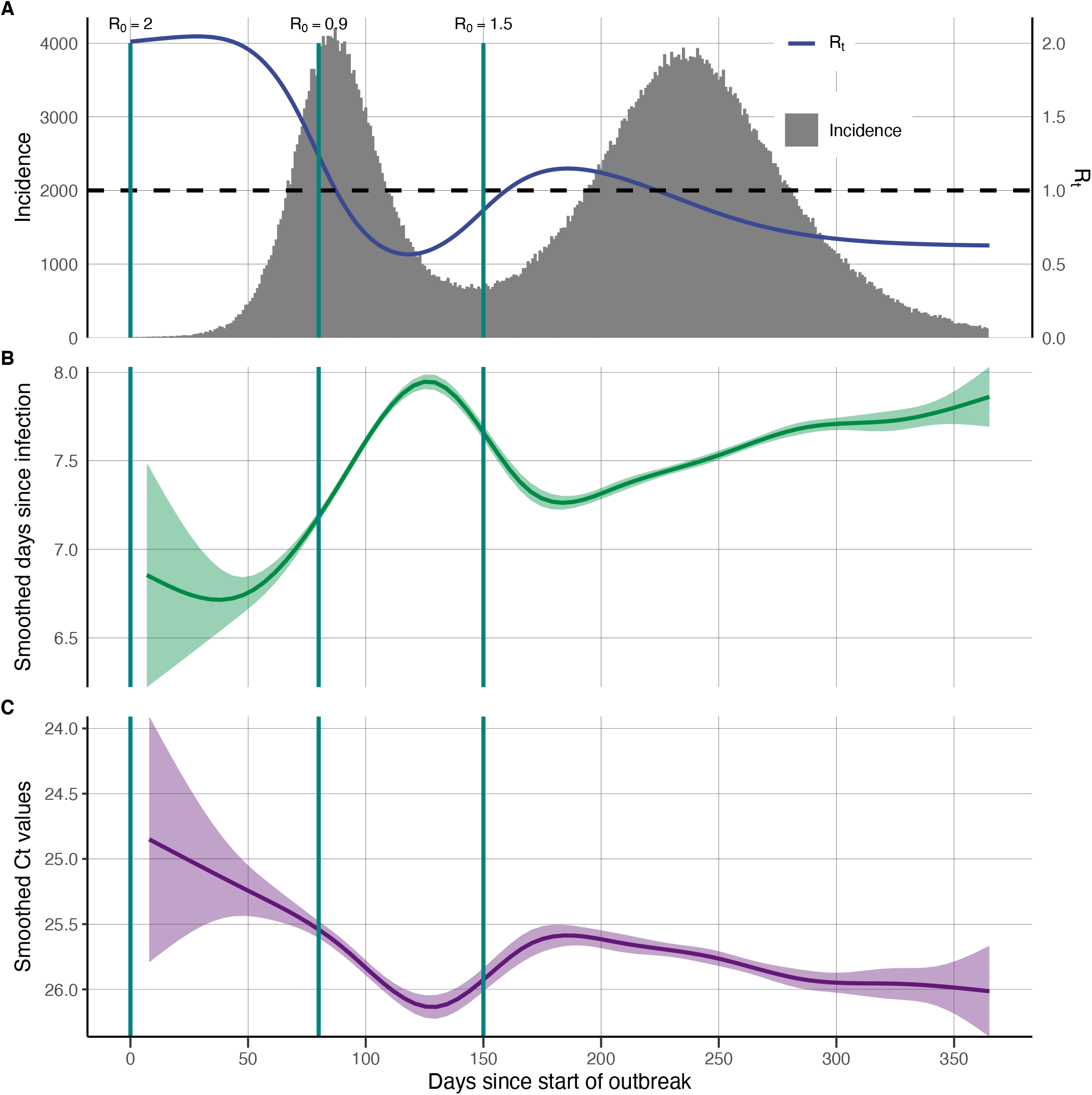

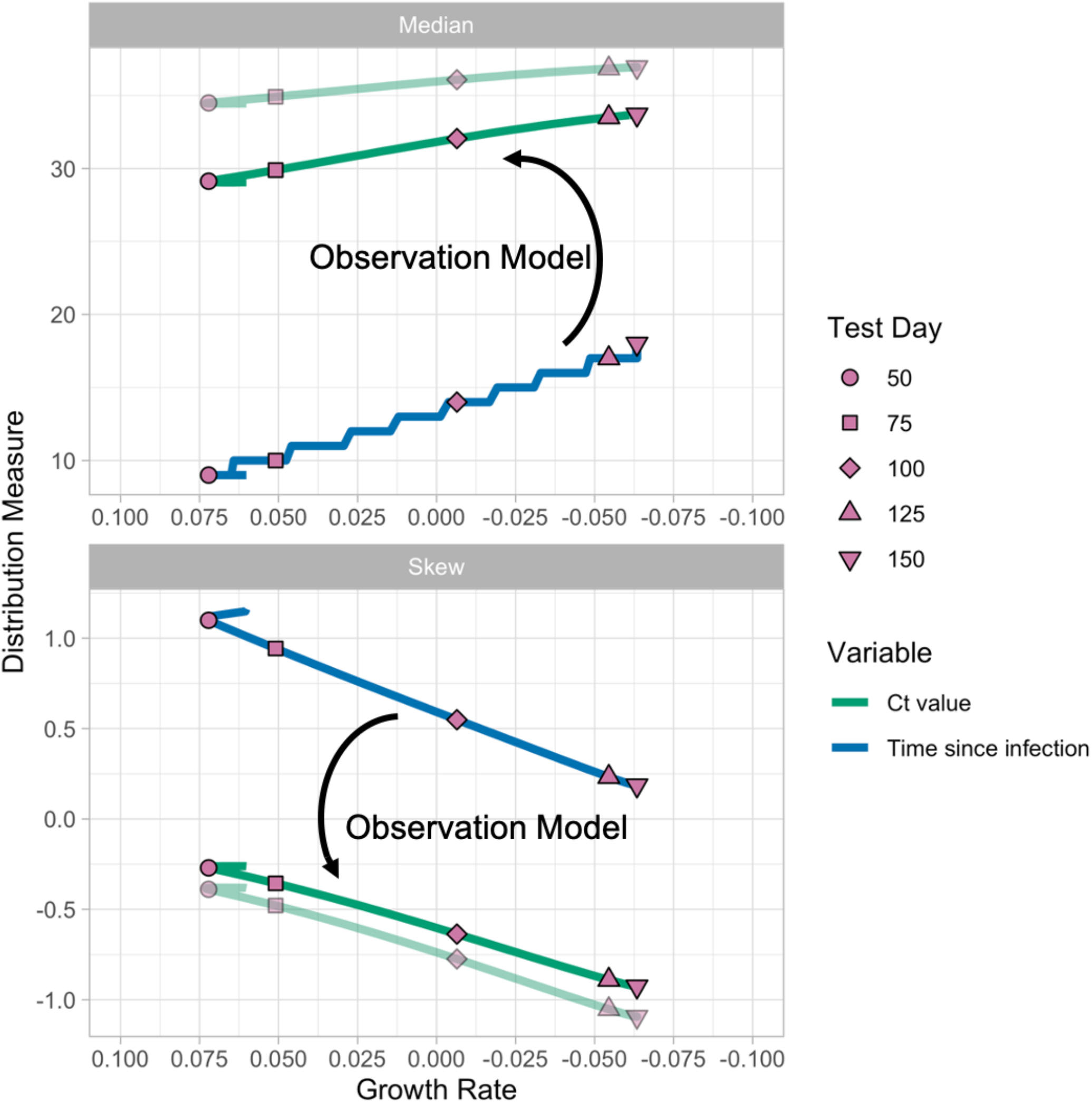

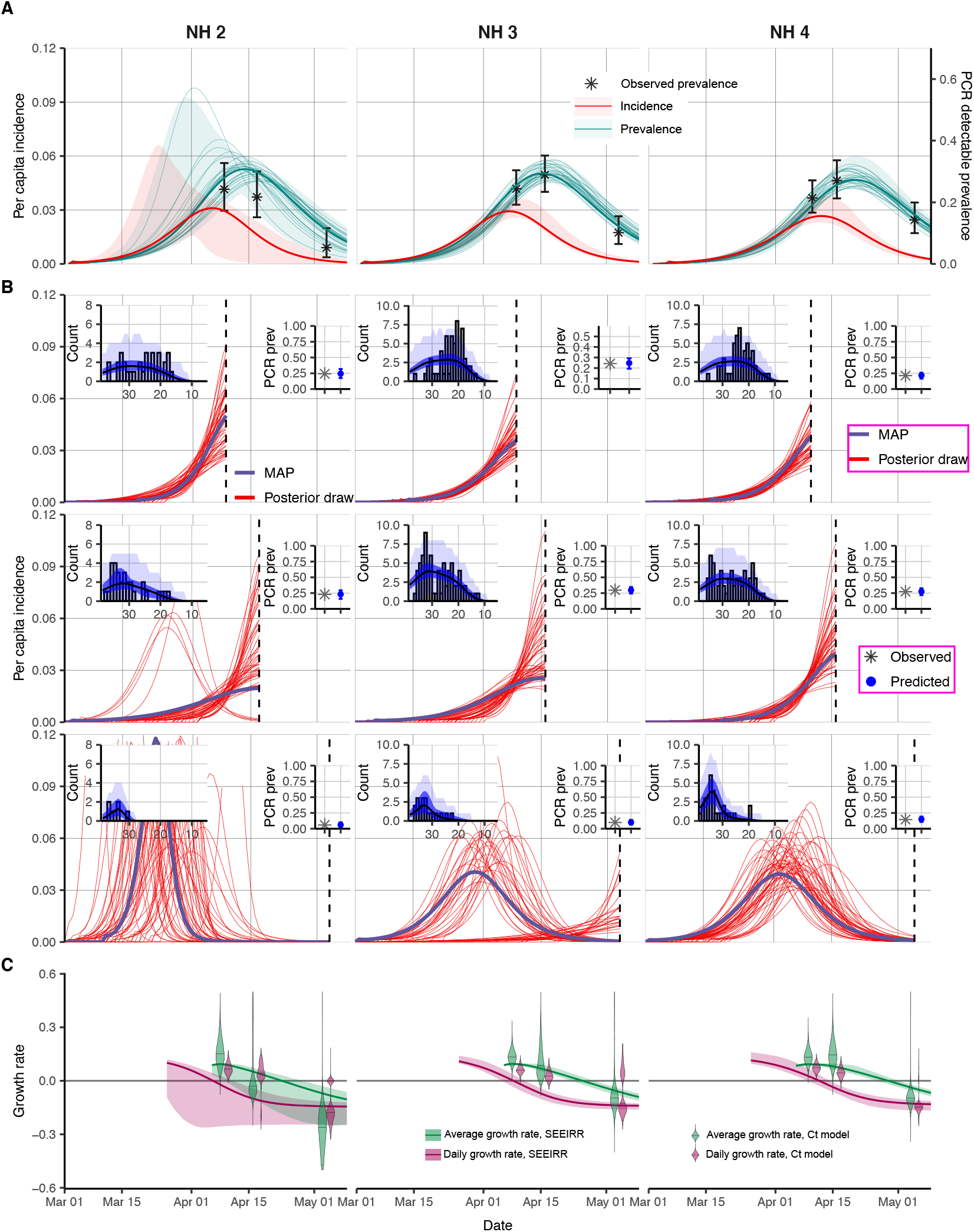

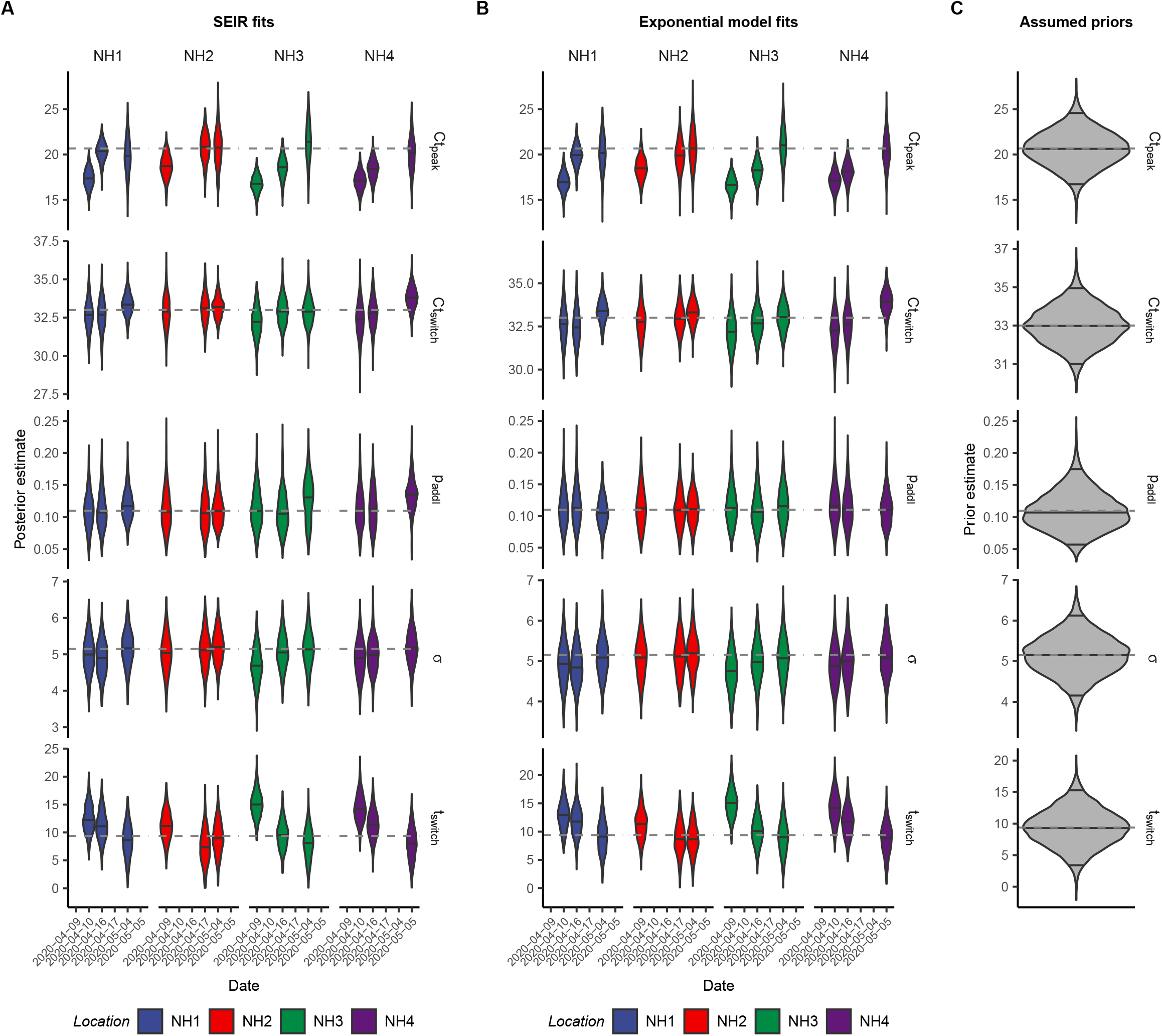

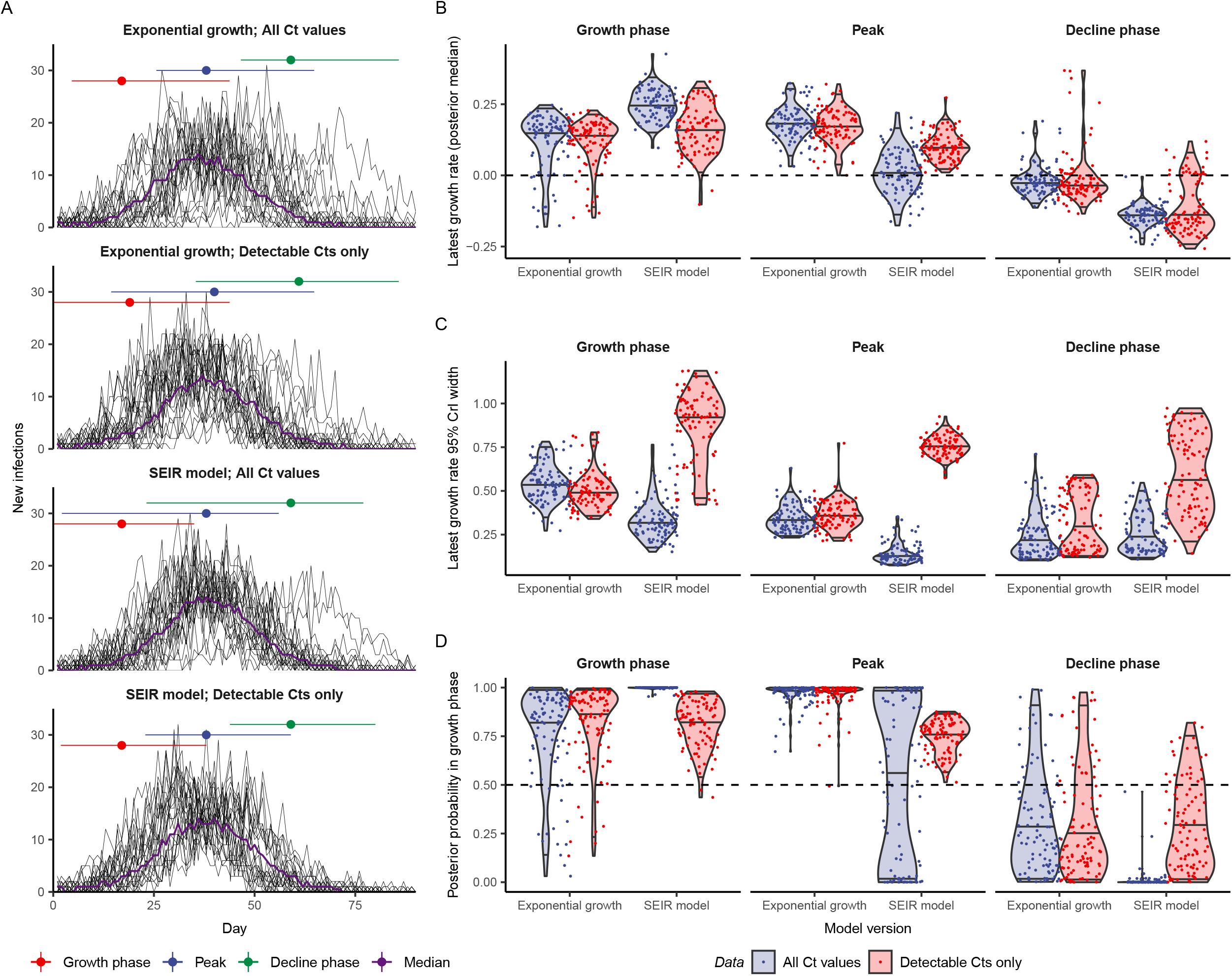

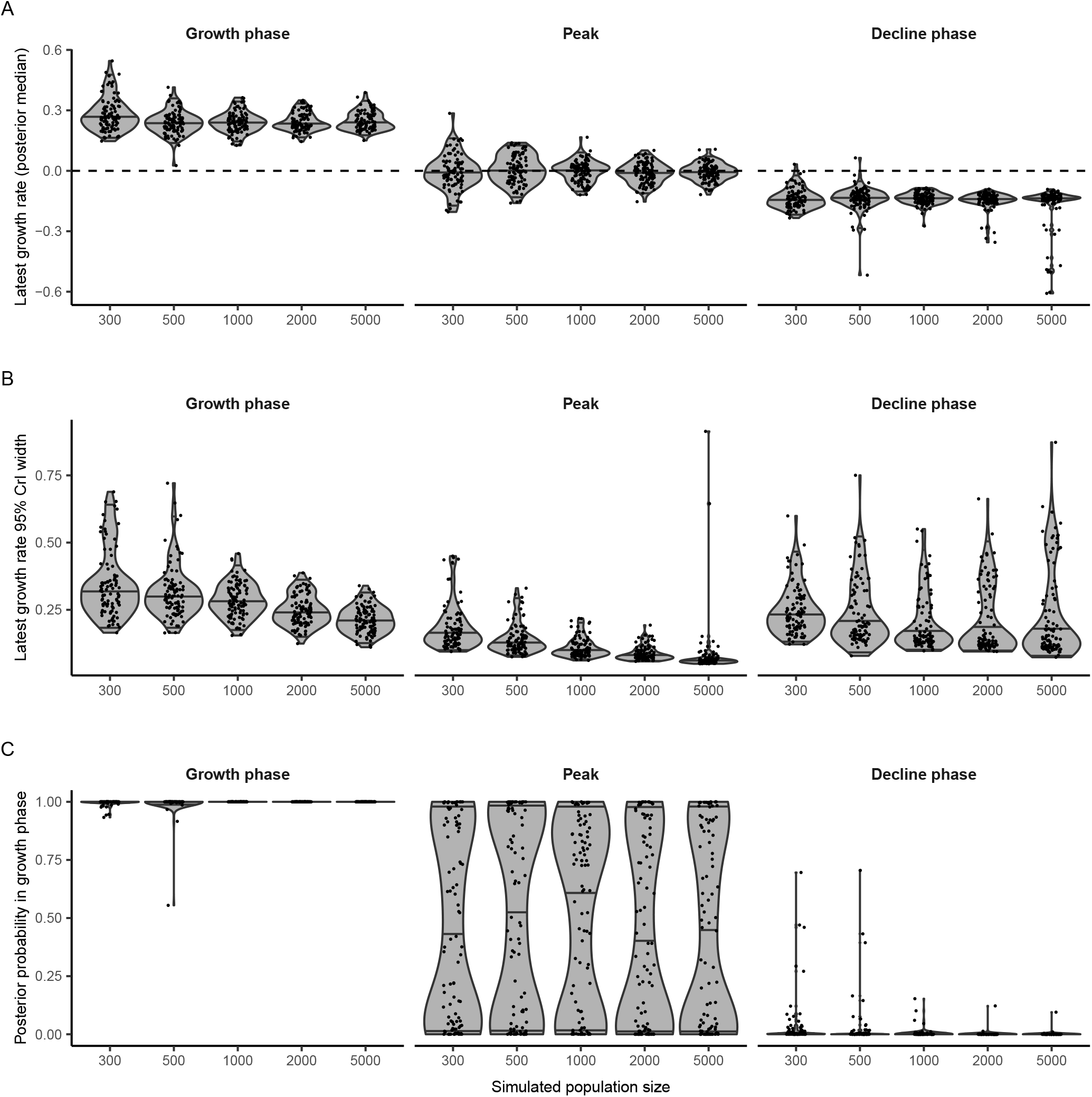

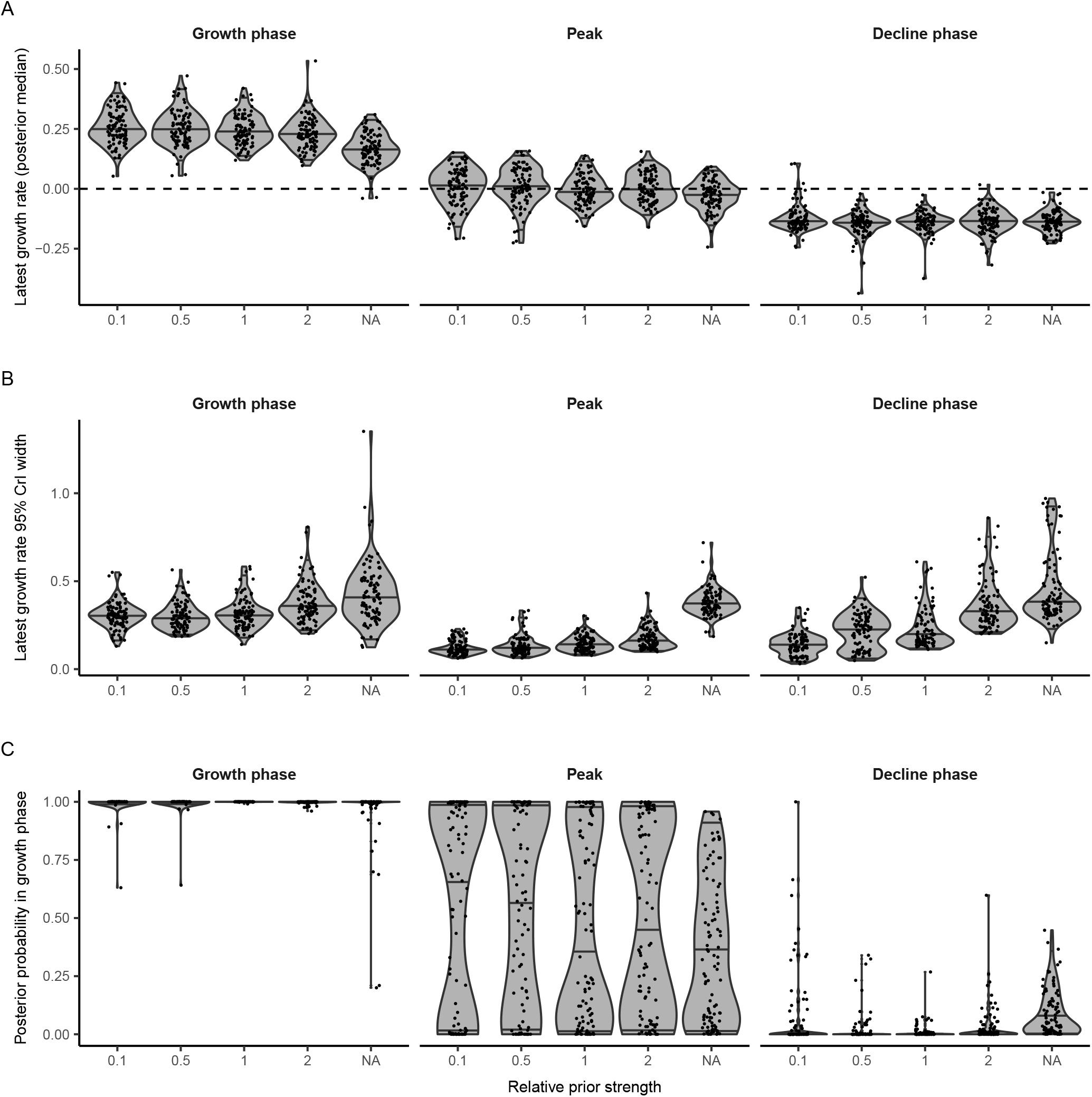

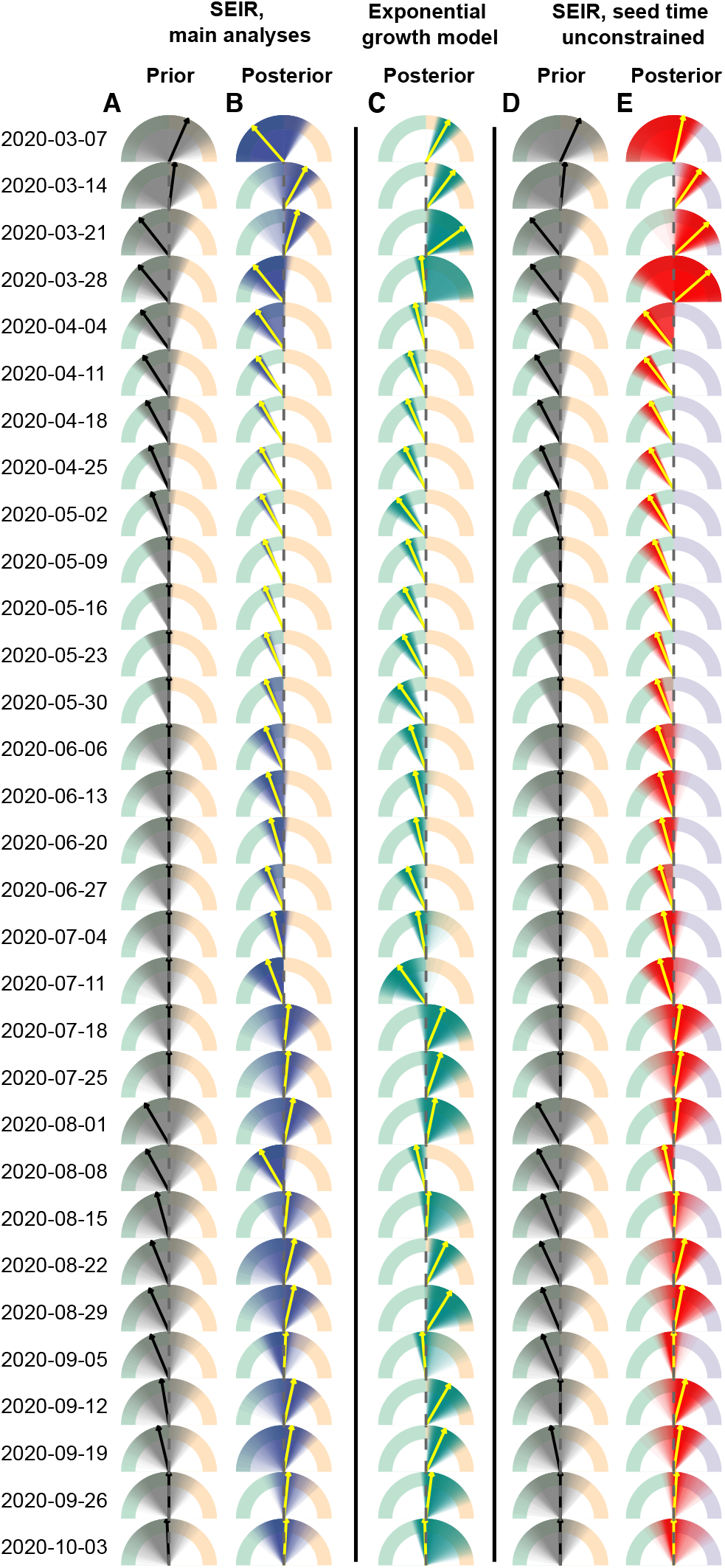

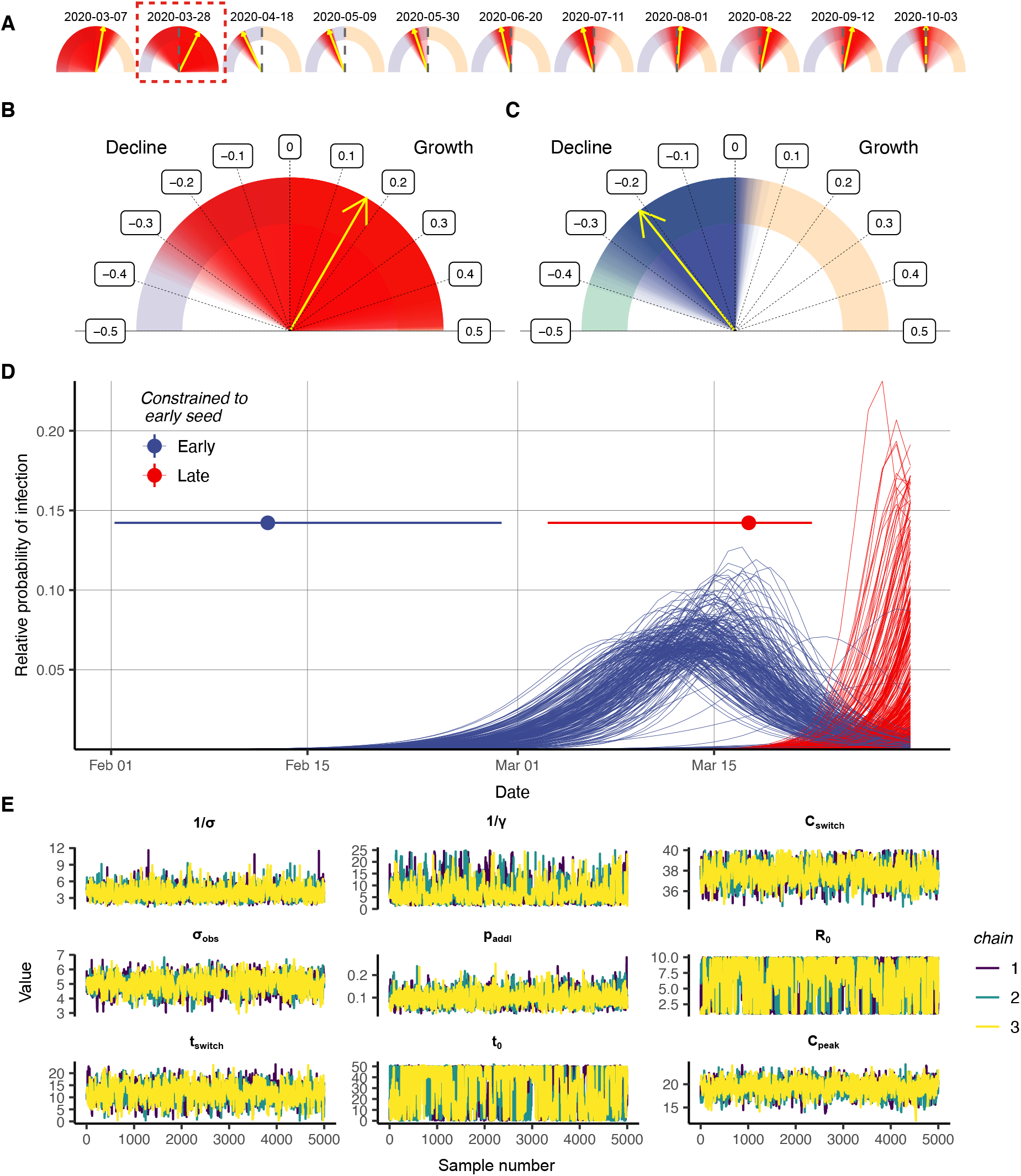

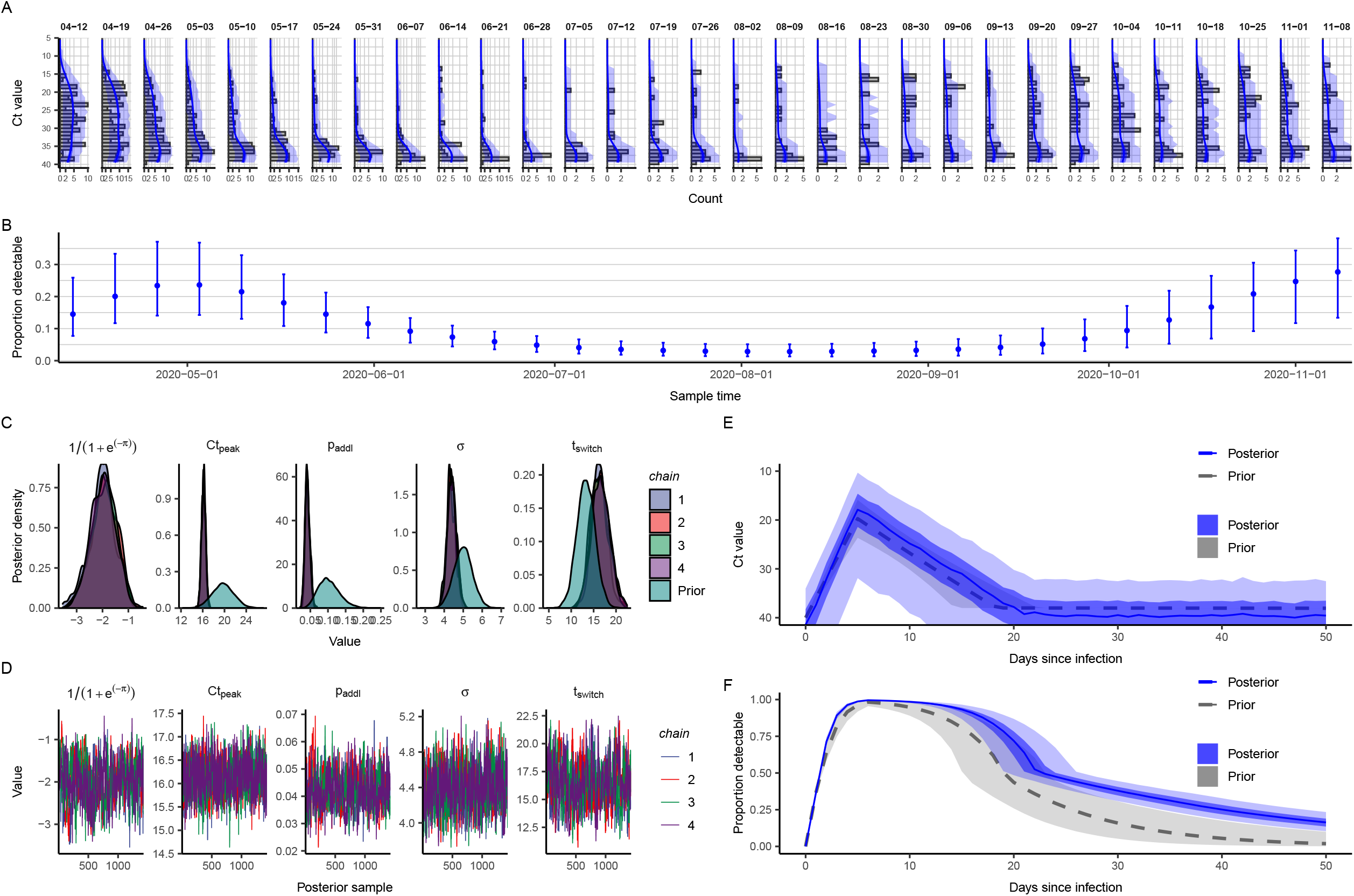

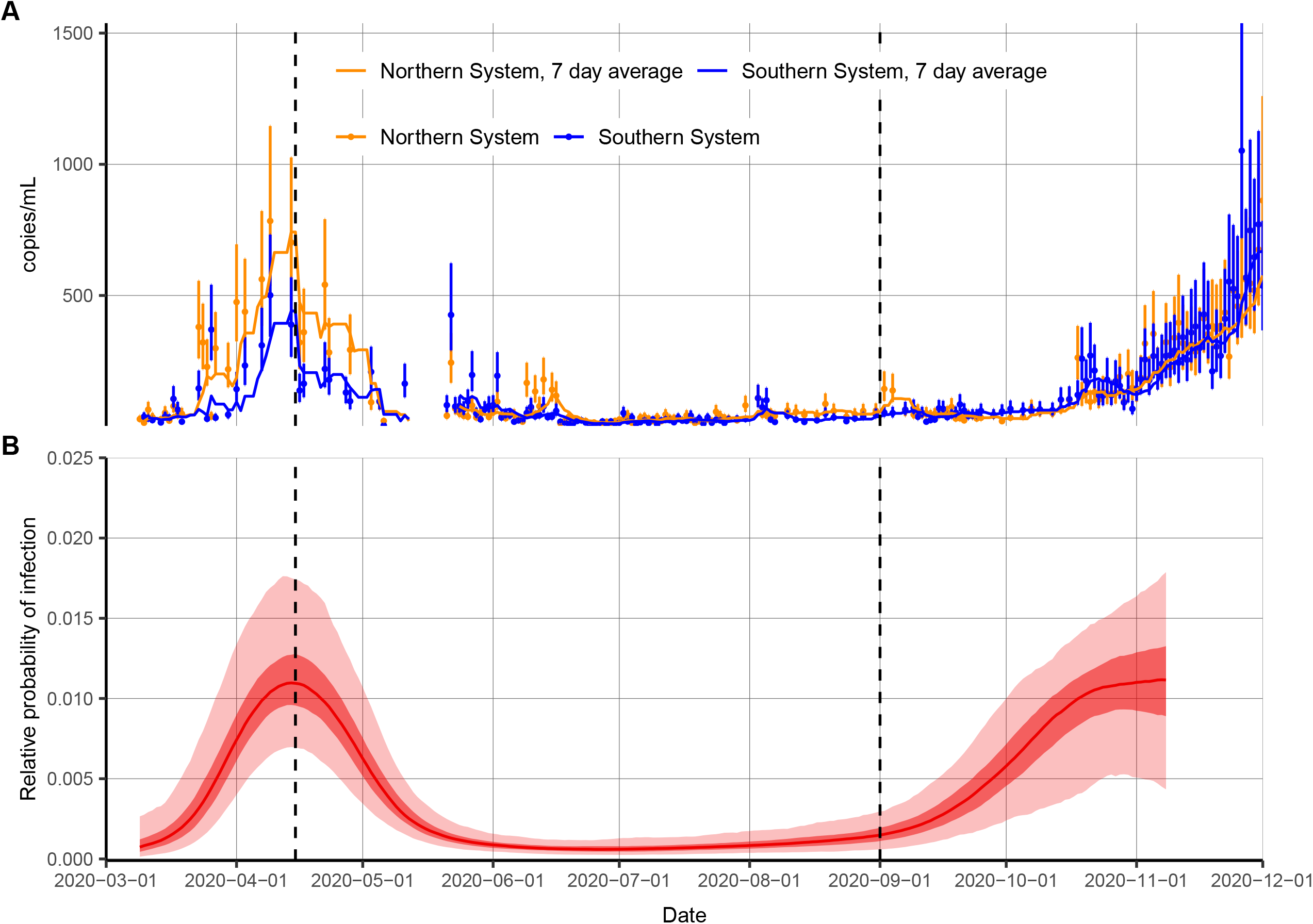

## Notes

### Funding Statement

This work is supported by U.S. National Institutes of Health Director’s Early Independence Award DP5-OD028145 (MJM, JAH), the Morris-Singer Fund (LKS and ML), U.S. Centers for Disease Control and Prevention Award U01IP001121 (LKS and ML), and U.S. National Institute of General Medical Sciences award U54GM088558 (ML, JAH, LKS).

### Author Declarations

IRB approval was obtained from Harvard School of Public Health for use of de-identified hospital-based viral load data (IRB 20-1703) and the Broad institute ORSP provided an exempt approval states for use of surveillance-based Ct values.

### Summary of Updates

Significant additions to the methodology, figures and analyses.

## References

1. H. V. Fineberg, M. E. Wilson. Epidemic science in real time. Science 324, 987 (2009).

2. World Health Organization. Public health surveillance for COVID-19: interim guidance. https://www.who.int/publications/i/item/who-2019-nCoV-surveillanceguidance-2020.8 (Accessed February 12, 2021).

3. T. Jombart, K. van Zandvoort, T. W. Russell, C. I. Jarvis, A. Gimma, S. Abbott, S. Clifford, S. Funk, H. Gibbs, Y. Liu, C. A. B. Pearson, N. I. Bosse, Centre for the Mathematical Modelling of Infectious Diseases COVID-19 Working Group, R. M. Eggo, A. J. Kucharski, W. J. Edmunds. Inferring the number of COVID-19 cases from recently reported deaths [version 1; peer review: 2 approved]. Wellcome Open Res. 5, 78 (2020).

4. M. Lipsitch, D. L. Swerdlow, L. Finelli. Defining the epidemiology of COVID-19—studies needed. NEJM 382, 1194–1196 (2020).

5. R. A. Betensky, Y. Feng. Accounting for incomplete testing in the estimation of epidemic parameters. Int. J. Epidemiol. 49, 1419–1426 (2020).

6. J. L. Vaerman, P. Saussoy, I. Ingargiola. Evaluation of real-time PCR data. J. Biol. Regul. Homeost. Agents 18, 212–214 (2004).

7. M. R. Tom, M. J. Mina. To interpret the SARS-CoV-2 test, consider the cycle threshold value. Clin. Inf. Dis. 71, 2252–2254 (2020).

8. T. C. Quinn, M. J. Wawer, N. Sewankambo, D. Serwadda, C. Li, F. Wabwire-Mangen, M. O. Meehan, T. Lutalo, R. H. Gray. Viral load and heterosexual transmission of Human Immunodeficiency Virus Type 1. NEJM 342, 921–929 (2000).

9. J. A. Fuller, M. K. Njenga, G. Bigogo, B. Aura, M. O. Ope, L. Nderitu, L. Wakhule, D. D. Erdman, R. F. Breiman, D. R. Feikin. Association of the CT values of real-time PCR of viral upper respiratory tract infection with clinical severity, Kenya. J. Med. Virol. 85, 924–932 (2013).

10. S. Bolotin, S. L. Deeks, A. Marchand-Austin, H. Rilkoff, V. Dang, R. Walton, A. Hashim, D. Farrell, N. S. Crowcroft. Correlation of Real Time PCR Cycle Threshold Cut-Off with Bordetella pertussis Clinical Severity. PLoS One 10, e0133209 (2015).

11. T. K. Tsang, B. J. Cowling, V. J. Fang, K. H. Chan, D. K. M. Ip, G. M. Leung, J. S. M. Peiris, S. Cauchemez. Influenza A virus shedding and infectivity in households. J. Inf. Dis. 212, 1420– 1428 (2015).

12. M. Moraz, D. Jacot, M. Papadimitriou-Olivgeris, L. Senn, G. Greub, K. Jaton, O. Opota. Universal admission screening strategy for COVID-19 highlighted the clinical importance of reporting SARS-CoV-2 viral loads. New Microbes New Infect. 38, 100820 (2020).

13. Y. Chen, L. Li. SARS-CoV-2: virus dynamics and host response. Lancet Inf. Dis. 20, 515–516 (2020).

14. D. Jacot, G. Greub, K. Jaton, O. Opota. Viral load of SARS-CoV-2 across patients and compared to other respiratory viruses. Microbes Infect. 22, 617–621 (2020).

15. K. A. Walsh, K. Jordan, B. Clyne, D. Rohde, L. Drummond, P. Byrne, S. Ahern, P. G. Carty, K. K. O’Brien, E. O’Murchu, M. O’Neill, S. M. Smith, M. Ryan, P. Harrington. SARS-CoV-2 detection, viral load and infectivity over the course of an infection. J. Infect. 81, 357–371 (2020).

16. A. S. Walker, E. Pritchard, T. House, J. V. Robotham, P. J. Birrell, I. Bell, J. I. Bell, J. N. Newton, J. Farrar, I. Diamond, R. Studley, J. Hay, K. D. Vihta, T. Peto, N. Stoesser, P. C. Matthews, D. W. Eyre, K. B. Pouwels, COVID-19 Infection Survey Team. Viral load in community SARS-CoV-2 cases varies widely and temporally. https://www.medrxiv.org/content/10.1101/2020.10.25.20219048v1 (2020).

17. M. Gousseff, P. Penot, L. Gallay, D. Batisse, N. Benech, K. Bouiller, R. Collarino, A. Conrad, D. Slama, C. Joseph, A. Lemaignen, F.-X. Lescure, B. Levy, M. Mahevas, B. Pozzetto, N. Vignier, B. Wyplosz, D. Salmon, F. Goehringer, E. Botelho-Nevers. Clinical recurrences of COVID-19 symptoms after recovery: viral lapse, reinfection or inflammatory rebound? J. Infect. 81, 816–817 (2020).

18. G. Rydevik, G. T. Innocent, G. Marion, R. S. Davidson, P. C. L. White, C. Billinis, P. Barrow, P. P. C. Mertens, D. Gavier-Widén, M. R. Hutchings. Using combined diagnostic test results to hindcast trends of infection from cross-sectional data. PLoS Comput. Biol. 12, e1004901 (2016).

19. J. Wallinga, M. Lipsitch. How generation intervals shape the relationship between growth rates and reproductive numbers. Proc. R. Soc. B 274, 599–604 (2007).

20. B. Borremans, N. Hens, P. Beutels, H. Leirs, J. Reijniers. Estimating time of infection using prior serological and individual information can greatly improve incidence estimation of human and wildlife infections. PLoS Comput. Biol. 12, e1004882 (2016).

21. H. Salje, D. A. T. Cummings, I. Rodriguez-Barraquer, L. C. Katzelnick, J. Lessler, C. Klungthong, B. Thaisomboonsuk, A. Nisalak, A. Weg, D. Ellison, L. Macareo, I. K. Yoon, R. Jarman, S. Thomas, A. L. Rothman, T. Endy, S. Cauchemez. Reconstruction of antibody dynamics and infection histories to evaluate dengue risk. Nature 557, 719–723 (2018).

22. J. A. Hay, A. Minter, K. E. C. Ainslie, J. Lessler, B. Yang, D. A. T. Cummings, A. J. Kucharski, S. Riley. An open source tool to infer epidemiological and immunological dynamics from serological data: serosolver. PLoS Comput. Biol. 16, e1007840 (2020).

23. H. E. de Melker, F. G. A. Versteegh, J. F. P. Schellekens, P. F. M. Teunis, M. Kretzschmar. The incidence of Bordetella pertussis infections estimated in the population from a combination of serological surveys. J. Infect. 53, 106–113 (2006).

24. J. Simonsen, K. Mølbak, G. Falkenhorst, K. A. Krogfelt, A. Linneberg, P. F. M. Teunis. Estimation of incidences of infectious diseases based on antibody measurements. Stat. Med. 28, 1882–1895 (2009).

25. P. F. M. Teunis, J. C. H. van Eijkeren, C. W. Ang, Y. T. H. P. van Duynhoven, J. B. Simonsen, M. A. Strid, W. van Pelt. Biomarker dynamics: estimating infection rates from serological data. Stat. Med. 31, 2240–2248 (2012).

26. D. A. Helb, K. K. A. Tetteh, P. L. Felgner, J. Skinner, A. Hubbard, E. Arinaitwe, H. Mayanja-Kizza, I. Ssewanyana, M. R. Kamya, J. G. Beeson, J. Tappero, D. L. Smith, P. D. Crompton, P. J. Rosenthal, G. Dorsey, C. J. Drakeley, B. Greenhouse. Novel serologic biomarkers provide accurate estimates of recent Plasmodium falciparum exposure for individuals and communities. Proc. Natl. Acad. Sci. U.S.A. 112, E4438–E4447 (2015).

27. K. M. Pepin, S. L. Kay, B. D. Golas, S. S. Shriner, A. T. Gilbert, R. S. Miller, A. L. Graham, S. Riley, P. C. Cross, M. D. Samuel, M. B. Hooten, J. A. Hoeting, J. O. Lloyd-Smith, C. T. Webb, M. G. Buhnerkempe. Inferring infection hazard in wildlife populations by linking data across individual and population scales. Ecol. Lett. 20, 275–292 (2017).

28. K. M. Gostic, L. McGough, E. B. Baskerville, S. Abbott, K. Joshi, C. Tedijanto, R. Kahn, R. Niehaus, J. A. Hay, P. M. De Salazar, J. Hellewell, S. Meakin, J. D. Munday, N. I. Bosse, K. Sherrat, R. N. Thompson, L. F. White, J. S. Huisman, J. Scire, S. Bonhoeffer, T. Stadler, J. Wallinga, S. Funk, M. Lipsitch, S. Cobey. Practical considerations for measuring the effective reproductive number, Rt. PLoS Comp. Biol. 16, e1008409 (2020).

29. N. J. Lennon, R. P. Bhattacharyya, M. J. Mina, H. L. Rehm, D. T. Hung, S. Smole, A. Woolley, E. S. Lander, S. B. Gabriel. Comparison of viral loads in individuals with or without symptoms at time of COVID-19 testing among 32,480 residents and staff of nursing homes and assisted living facilities in Massachusetts. https://www.medrxiv.org/content/10.1101/2020.07.20.20157792v1 (2020).

30. T. K. Tsang, P. Wu, Y. Lin, E. H. Y. Lau, G. M. Leung, B. J. Cowling. Effect of changing case definitions for COVID-19 on the epidemic curve and transmission parameters in mainland China: a modelling study. Lancet Public Health 5, e289–296 (2020).

31. S. Abbott, J. Hellewell, R. N. Thompson, K. Sherratt, H. P. Gibbs, N. I. Bosse, J. D. Munday, S. Meakin, E. L. Doughty, J. Y. Chun, Y. W. D. Chan, F. Finger, P. Campbell, A. Endo, C. A. B. Pearson, A. Gimma, T. Russell, CMMID COVID modelling group, S. Flasche, A. J. Kucharski, R. M. Eggo, S. Funk. Estimating the time-varying reproduction number of SARS-CoV-2 using national and subnational case counts [version 2; peer review: awaiting peer review]. Wellcome Open Res. 5, 112 (2020).

32. X. Xu, T. Kypraios, P.D. O’Neill. Bayesian non-parametric inference for stochastic epidemic models using Gaussian processes. Biostatistics 17, 619–633 (2016).

33. Massachusetts Water Resources Authority. Wastewater COVID-19 Tracking. https://www.mwra.com/biobot/biobotdata.htm (Accessed January 7, 2021).

34. S. Riley, C. Atchison, D. Ashby, C. A. Donnelly, W. Barclay, G. Cooke, H. Ward, A. Darzi, P. Elliott, REACT Study Group. REal-time Assessment of Community Transmission (REACT) of SARS-CoV-2 virus: study protocol [version 1; peer review: 1 approved, 1 approved with reservations]. Wellcome Open Res. 5, 200 (2020).

35. R. Niehus, E. van Kleef, A. Turlej-Rogacka, C. Lammens, Y. Carmeli, H. Goossens, E. Tacconelli, B. Carevic, L. Preotescu, S. Malhotra-Kumar, B. S. Cooper. Quantifying antibiotic impact on within-patient dynamics of extended-spectrum beta-lactamase resistance. eLife 9, e49206 (2020).

36. D. Rhoads, D. R. Peaper, R. C. She, F. S. Nolte, C. M. Wojewoda, N. W. Anderson, B. S. Pritt. College of American Pathologists (CAP) Microbiology Committee perspective: caution must be used in interpreting the cycle threshold (Ct) value. Clin. Infect. Dis. 10.1093/cid/ciaa1199 (2020).

37. A. T. Xiao, Y. X. Tong, S. Zhang. Profile of RT-PCR for SARS-CoV-2: a preliminary study from 56 COVID-19 patients. Clin. Inf. Dis. 71, 2249–2251 (2020).

38. W. C. Ko, J. M. Rolain, N. Y. Lee, P. L. Chen, C. T. Huang, P. I. Lee, P. R. Hsueh. Arguments in favour of remdesivir for treating SARS-CoV-2 infections. Int. J. Antimicrob. Agents 55, 105933 (2020).

39. E. Mahase. COVID-19: what have we learnt about the new variant in the UK? BMJ 371, m4944 (2020).

40. D. B. Larremore, B. Wilder, E. Lester, S. Shehata, J. M. Burke, J. A. Hay, M. Tambe, M. J. Mina, R. Parker. Test sensitivity is secondary to frequency and turnaround time for COVID-19 screening. Sci. Adv. 7, eabd5393 (2021).

41. A. Tahamtan, A. Ardebili. Real-time RT-PCR in COVID-19 detection: issues affecting the results. Expert Rev. Mol. Diagn. 20, 453–454 (2020).

42. K. K. W. To, O. T. Y. Tsang, W. S. Leung, A. R. Tam, T. C. Wu, D. C. Lung, C. C. Y. Yip, J. P. Cai, J. M. C. Chan, T. S. H. Chik, D. P. L. Lau, C. Y. C. Choi, L. L. Chen, W. M. Chan, K. H. Chan, J. D. Ip, A. C. K. Ng, R. W. S. Poon, C. T. Luo, Vi. C. C. Cheng, J. F. W. Chan, I. F. N. Hung, Z. Chen, H. Chen, K. Y. Yuen. Temporal profiles of viral load in posterior oropharyngeal saliva samples and serum antibody responses during infection by SARS-CoV-2: an observational cohort study. Lancet Inf. Dis. 20, 565–574 (2020).

43. R. Wölfel, V. M. Corman, W. Guggemos, M. Seilmaier, S. Zange, M. A. Müller, D. Niemeyer, T. C. Jones, P. Vollmar, C. Rothe, M. Hoelscher, T. Bleicker, S. Brünink, J. Schneider, R. Ehmann, K. Zwirglmaier, C. Drosten, C. Wendtner. Virological assessment of hospitalized patients with COVID-19. Nature 581, 465–469 (2020).

44. Q. Z. Long, X. J. Tang, Q. L. Shi, Q. Li, H. J. Deng, J. Yuan, J. L. Hu, W. Xu, Y. Zhang, F. J. Lv, K. Su, F. Zhang, J. Gong, B. Wu, X. M. Liu, J. J. Li, J. F. Qiu, J. Chen, A. L. Huang. Clinical and immunological assessment of asymptomatic SARS-CoV-2 infections. Nature Med. 26, 1200–1204 (2020).

45. Y. Liu, L. M. Yan, L. Wan, T. X. Xiang, A. Le, J. M. Liu, M. Peiris, L. L. M. Poon, W. Zhang. Viral dynamics in mild and severe cases of COVID-19. Lancet Inf. Dis. 20, 656–657 (2020).

46. A. Chandrashekar, J. Liu, A. J. Martinot, K. McMahan, N. B. Mercado, L. Peter, L. H. Tostanoski, J. Yu, Z. Maliga, M. Nekorchuk, K. Busman-Sahay, M. Terry, L. M. Wrijil, S. Ducat, D. R. Martinez, C. Atyeo, S. Fischinger, J. S. Burke, M. D. Slein, L. Pessaint, A. Van Ry, J. Greenhouse, T. Taylor, K. Blade, A. Cook, B. Finneyfrock, R. Brown, E. Teow, J. Velasco, R. Zahn, F. Wegmann, P. Abbink, E. A. Bondzie, G. Dagotto, M. S. Gebre, X. He, C. Jacob-Dolan, N. Kordana, Z. Li, M. A. Lifton, S. H. Mahrokhian, L. F. Maxfield, R. Nityanandam, J. P. Nkolola, A. G. Schmidt, A. D. Miller, R. S. Baric, G. Alter, P. K. Sorger, J. D. Estes, H. Andersen, M. G. Lewis, D. H. Barouch. SARS-CoV-2 infection protects against rechallenge in rhesus macaques. Science 369, 812–817 (2020).

47. J. Yu, L. H. Tostanoski, L. Peter, N. B. Mercado, K. McMahan, S. H. Mahrokhian, J. P. Nkolola, J. Liu, Z. Li, A. Chandrashekar, D. R. Martinez, C. Loos, C. Atyeo, S. Fischinger, J. S. Burke, M. D. Slein, Y. Chen, A. Zuiani, F. J. N. Lelis, M. Travers, S. Habibi, L. Pessaint, A. Van Ry, K. Blade, R. Brown, A. Cook, B. Finneyfrock, A. Dodson, E. Teow, J. Velasco, R. Zahn, F. Wegmann, E. A. Bondzie, G. Dagotto, M. S. Gebre, X. He, C. Jacob-Dolan, M. Kirilova, N. Kordana, Z. Lin, L. F. Maxfield, F. Nampanya, R. Nityanandam, J. D. Ventura, H. Wan, Y. Cai, B. Chen, A. G. Schmidt, D. R. Wesemann, R. S. Baric, G. Alter, H. Andersen, M. G. Lewis, D. H. Barouch. DNA vaccine protection against SARS-CoV-2 in rhesus macaques. Science 369, 806–811 (2020).

48. S. F. Sia, L. M. Yan, A. W. H. Chin, K. Fung, K. T. Choy, A. Y. L. Wong, P. Kaewpreedee, R. A. P. M. Perera, L. L. M. Poon, J. M. Nicholls, M. Peiris, H. L. Yen. Pathogenesis and transmission of SARS-CoV-2 in golden hamsters. Nature 583, 834–838 (2020).

49. A. M. Bosco-Lauth, A. E. Hartwig, S. M. Porter, P. W. Gordy, M. Nehring, A. D. Byas, S. VandeWoude, I. K. Ragan, R. M. Maison, R. A. Bowen. Experimental infection of domestic dogs and cats with SARS-CoV-2: pathogenesis, transmission, and response to reexposure in cats. Proc. Natl. Acad. Sci. U.S.A. 117, 26382–26388 (2020).

50. M. M. Arons, K. M. Hatfield, S. C. Reddy, A. Kimball, A. James, J. R. Jacobs, J. Taylor, K. Spicer, A. C. Bardossy, L. P. Oakley, S. Tanwar, J. W. Dyal, J. Harney, Z. Chisty, J. M. Bell, M. Methner, P. Paul, C. M. Carlson, H. P. McLaughlin, N. Thornburg, S. Tong, A. Tamin, Y. Tao, A. Uehara, J. Harcourt, S. Clark, C. Brostrom-Smith, L. C. Page, M. Kay, J. Lewis, P. Montgomery, N. D. Stone, T. A. Clark, M. A. Honein, J. S. Duchin, J. A. Jernigan. Presymptomatic SARS-CoV-2 infections and transmission in a skilled nursing facility. NEJM 382, 2081–2090 (2020).

51. L. Ferretti, C. Wymant, M. Kendall, L. Zhao, A. Nurtay, L. Abeler-Dörner, M. Parker, D. Bonsall, C. Fraser. Quantifying SARS-CoV-2 transmission suggests epidemic control with digital contact tracing. Science 368, eabb6936 (2020).

52. S. A. Lauer, K. H. Grantz, Q. Bi, F. K. Jones, Q. Zheng, H. R. Meredith, A. S. Azman, N. G. Reich, J. Lessler. The incubation period of coronavirus disease 2019 (COVID-19) from publicly reported confirmed cases: estimation and application. Ann. Int. Med. 172, 577–582 (2020).

53. X. He, E. H. Y. Lau, P. Wu, X. Deng, J. Wang, X. Hao, Y. C. Lau, J. Y. Wong, Y. Guan, X. Tan, X. Mo, Y. Chen, B. Liao, W. Chen, F. Hu, Q. Zhang, M. Zhong, Y. Wu, L. Zhao, F. Zhang, B. J. Cowling, F. Li, G. M. Leung. Temporal dynamics in viral shedding and transmissibility of COVID-19. Nature Med. 26, 672–675 (2020).

54. M. Cevik, M. Tate, O. Lloyd, A. E. Maraolo, J Schafers, A. Ho. SARS-CoV-2, SARS-CoV, and MERS-CoV viral load dynamics, duration of viral shedding and infectiousness: a systematic review and meta-analysis. Lancet Microbe 2, e13–e22 (2021).

55. B. Borremans, A. Gamble, K. C. Prager, S. K. Helman, A. M. McClain, C. Cox, V. Savage, J. O. Lloyd-Smith. Quantifying antibody kinetics and RNA detection during early-phase SARS-CoV-2 infection by time since symptom onset. eLife 9, e60122 (2020).

56. A. Singanayagam, M. Patel, A. Charlett, J. L. Bernal, V. Saliba, J. Ellis, S. Ladhani, M. Zambon, R. Gopal. Duration of infectiousness and correlation with RT-PCR cycle threshold values in cases of COVID-19, England, January to May 2020. Euro Surveill. 25, 2001483 (2020).

57. B. La Scola, M. Le Bideau, J. Andreani, V. T. Hoang, C. Grimaldier, P. Colson, P. Gautret, D. Raoult. Viral RNA load as determined by cell culture as a management tool for discharge of SARS-CoV-2 patients from infectious disease wards. Eur. J. Clin. Microbiol. Infect. Dis. 39, 1059–1061 (2020).

58. J. Bullard, K. Dust, D. Funk, J. E. Strong, D. Alexander, L. Garnett, C. Boodman, A. Bello, A. Hedley, Z. Schiffman, K. Doan, N. Bastien, Y. Li, P. G. Van Caeseele, G. Poliquin. Clin. Infect. Dis. 71, 2663–2666 (2020).

59. J. van Beek, Z. Igloi, T. Boelsums, E. Fanoy, H. Gotz, R. Molenkamp, J. van Kampen, C. GeurtsvanKessel, A. van der Eijk, D. van de Vijver, M. Koopmans. From more testing to smart testing: data-guided SARS-CoV-2 testing choices. https://www.medrxiv.org/content/10.1101/2020.10.13.20211524v2 (2020).

60. J. Y. Kim, J. H. Ko, Y. Kim, Y. J. Kim, J. M. Kim, Y. S. Chung, H. M. Kim, M. G. Han, S. Y. Kim, B. S. Chin. Viral load kinetics of SARS-CoV-2 infection in first two patients in Korea. J. Korean Med. Sci. 35, e86 (2020).

61. N. Li, X. Wang, T. Lv. Prolonged SARS-CoV-2 RNA shedding: not a rare phenomenon. J. Med. Virol. 92, 2286–2287 (2020).

62. H. Kawasuji, Y. Takegoshi, M. Kaneda, A. Ueno, Y. Miyajima, K. Kawago, Y. Fukui, Y. Yoshida, M. Kimura, H. Yamada, I. Sakamaki, H. Tani, Y. Morinaga, Y. Yamamoto. Viral load dynamics in transmissible symptomatic patients with COVID-19. https://www.medrxiv.org/content/10.1101/2020.06.02.20120014v1 (2020).

63. D. J. Earl and M. W. Deem. Parallel Tempering: Theory, Applications, and New Perspectives. Phys. Chem. Chem. Phys. 7, 3910–3916 (2005).

64. M. Plummer, N. Best, K. Cowles, K. Vines. CODA: Convergence Diagnosis and Output Analysis for MCMC. R News 6, 7–11 (2006).

65. R. McElreath. Statistical Rethinking: A Bayesian Course with Examples in R and STAN (Chapman and Hall/CRC Press, Boca Raton, FL, ed. 2, 2020).

