## Supplementary Methods and Figures for "Estimating epidemiologic dynamics from cross-sectional viral load distributions"

### Materials and Methods

Nursing Home Data

Data comes from nasopharyngeal specimens processed at the Broad Institute of MIT and Harvard CRSP CLIA laboratory, with an FDA Emergency Use Authorized laboratory developed assay. Ct values were provided along with a random tube ID and a unique anonymized institute ID to reflect that specimens came from distinct institutions. The specimens used here originated in early 2020 when public health efforts in MA led to comprehensively serial testing senior nursing facilities (*29*). Swabs from those public health efforts were processed for clinical diagnostics. The anonymized Ct data was made available and used for these analyses.

Brigham & Women's Hospital Data

Data comes from nasopharyngeal specimens processed on a Hologic Panther Fusion SARS-CoV-2 assay for patients at the Brigham & Women's Hospital in Boston, MA. Testing during the first two weeks in April 2020 were restricted to patients with symptoms consistent with COVID-19 and who needed hospital admission. Following April 15, testing criteria for this platform were expanded to include all asymptomatic hospital admissions, symptomatic patients in the emergency room who were not admitted to the hospital, and inpatients requiring testing who were not in labor. Symptomatic ER patients who were admitted to the hospital were tested on a different PCR platform. While this is not a perfectly representative surveillance sample, the routine testing of hospital admissions who were not seeking COVID treatment creates a cohort that is less biased than symptom-based testing and represents the overall rise and fall of cases in the hospital's catchment area. Daily data is aggregated by week. Daily confirmed case counts for Massachusetts were obtained from the NYT GitHub page (https://github.com/nytimes/covid-19-data).

Simulated Epidemic Transmission Models

We developed two compartmental models to describe SARS-CoV-2 transmission over the course of an epidemic. First, we developed a classical SEIR model with compartments for susceptible ($S$), exposed-not-infectious ($E$), infectious ($I$) and recovered ($R$). This is the model we use to generate Fig. 1 and to estimate incidence based on Ct distributions as shown in Fig. 2C. Note that this simpler model does not need to account for PCR detectability, as the time course of viral loads is modelled separately. The rationale behind parameter choices is described below. The compartmental transition equations are given by:

$$\frac{dS}{dt} = \frac{-\beta SI}{N},$$
$$\frac{dE}{dt} = \frac{\beta SI}{N} - \sigma E,$$
$$\frac{dI}{dt} = \sigma E - \gamma I, \text{ and}$$
$$\frac{dR}{dt} = \gamma I,$$

where $\beta = \frac{R_0}{\gamma}$, and $\beta = 0$ for $t < t_0$ .

Second, to describe the prevalence of individuals who are PCR positive for SARS-CoV-2, we developed an SEEIRR transmission model with states for susceptible (*S*), exposed-not-infectious-undetectable ($E_1$), exposed-not-infectious-detectable ($E_2$), infected-infectious-detectable (*I*), recovered-not-infectious-detectable ($R_1$) and recovered-not-infectious-undetectable ($R_2$), as depicted in Fig. S1A. These additional exposed and recovered compartments account for the periods when an individual will test PCR positive, but is not yet or is no longer infectious. We fit this model to data from the four Massachusetts nursing homes using Markov chain Monte Carlo (MCMC) to generate the trajectories in Fig. 2A, placing informative priors on the shared transition rate parameters (Tbl. S1) and uniform priors on the nursing home-specific basic reproductive numbers, $R_0$, and the effective seed times, $t_0$. The compartmental transition equattions are given by:

$$\frac{dS}{dt} = \frac{-\beta_l SI}{N},$$
$$\frac{dE_1}{dt} = \frac{\beta_l SI}{N} - \sigma' E,$$
$$\frac{dE_2}{dt} = \sigma' E_1 + \alpha E_2,$$
$$\frac{dI}{dt} = \alpha E_2 - \gamma' I,$$
$$\frac{dR_1}{dt} = \gamma' I - \omega R_1, \text{ and}$$
$$\frac{dR_2}{dt} = \omega R_1,$$

where $\beta_l$ is the location specific transmission rate and $\beta_l = 0$ for $t < t_{0,l}$ , the location specific seed time.

<u>Ct Value Model</u>

We developed a mathematical model describing the distribution of observed SARS-CoV-2 viral loads over time following infection. This model is similar to that used by Larremore et al.

(*40*) but allowing for more flexibility in the decline of viral load. We used a parametric model describing the modal Ct value, $C_{mode}(a)$, for an individual $a$ days after infection, represented by the solid black line in Fig. S1B. The measured Ct value is a linear function of the log of the viral load in the sample, but we describe the model on the Ct scale to match the data. Because we are interested in the population-level distribution and not individual trajectories, we assumed that observed Ct values $a$ days after infection followed a Gumbel distribution with location (mode) parameter $C_{mode}(a)$ and scale parameter $\sigma(a)$ that also may depend on the number of days $a$ after infection. Note that for the Gumbel distribution, the mean and variance are given by $C_{mode}(a) - \sigma(a)\gamma$ and $\frac{\sigma(a)^2\pi^2}{6}$, respectively, where $\gamma$ is Euler's constant. We chose a Gumbel distribution to capture overdispersion of high measured Ct values. This distribution captures the variation resulting from both swabbing variability and individual-level differences in viral kinetics. We note that at any point in the infection, there is a considerable amount of person-to-person and swab-to-swab variation in viral loads (*41–43*), including a possible difference by symptom status (*15, 44, 45*). Tracking individual-level viral kinetics would require a hierarchical model capturing individual-level parameters, but is not necessary for this analysis.

The modal Ct value at day $a$ follows a two-hinge function that is at the true undetectable value of $C_{zero}$ for $a \le t_{eclipse}$, decreases linearly (log viral load increases) to a minimum Ct of $C_{peak}$ at $a = t_{eclipse} + t_{peak}$, increases linearly (log viral load wanes) to a Ct value of $C_{switch}$ at $a = t_{eclipse} + t_{peak} + t_{switch}$, and then increases (log viral load wanes) at a slower linear rate until it reaches the limit of detection (LOD), $C_{LOD}$, at $a = t_{LOD}$. That is, the modal Ct value is given by:

$$C_{mode}(a) = \begin{cases} C_{zero}, & a \le t_{eclipse} \\ C_{zero} + \frac{C_{peak} - C_{zero}}{t_{peak}}(a - t_{eclipse}), & t_{eclipse} < a \le t_{eclipse} + t_{peak} \\ C_{peak} + \frac{C_{switch} - C_{peak}}{t_{switch}}(a - t_{eclipse} - t_{peak}), & t_{eclipse} + t_{peak} < a \le t_{eclipse} + t_{peak} + t_{switch} \\ C_{switch} + \frac{C_{LOD} - C_{switch}}{t_{LOD} - t_{switch} - t_{peak} - t_{eclipse}}(a - t_{eclipse} - t_{peak} - t_{switch}), & t_{eclipse} + t_{peak} + t_{switch} < a \end{cases}$$

The Ct value of a randomly-chosen individual $a$ days after infection, $C(a)$, is then distributed according to: $C(a) \sim Gumbel(C_{mode}(a), \sigma(a))$, where $\sigma(a)$ is given by:

$$\sigma(a) = \begin{cases} \sigma_{obs}, & a < t_{eclipse} + t_{peak} + t_{switch} \\ \sigma_{obs}\left[1 - \frac{1 - s_{mod}}{t_{mod}}(a - t_{eclipse} - t_{peak} - t_{switch})\right], & t_{eclipse} + t_{peak} + t_{switch} \le a < t_{eclipse} + t_{peak} + t_{switch} + t_{mod} \\ \sigma_{obs}s_{mod}, & t_{eclipse} + t_{peak} + t_{switch} + t_{mod} \le a \end{cases}$$

This observation model allows for Ct values to gradually shrink towards the mode as the infection is cleared and most individuals become undetectable again. The log-viral load (in $\log_{10}$ RNA copies per mL) $a$ days after infection, $V(a)$, can then be calculated by: $V(a) = V_{LOD} + \frac{C_{LOD} - C(a)}{\log_2(10)}$, where the limit of detection on the Ct scale is $C_{LOD} = 40$ and on the viral load scale depends on test characteristics (we assume $V_{LOD}$=3 $\log_{10}$ RNA copies per mL). This model captures the shape of the observed modal viral load over time and the features described above.

A feature of viral loads not captured by the above model is that a small fraction of individuals remain PCR positive at very high Ct values for many weeks after recovery, whereas most drop to undetectable levels within a couple of weeks. To account for this, each day after $t_{eclipse} + t_{peak} + t_{switch}$ there is a daily probability, $p_{addl}$, of an individual fully clearing the virus and becoming undetectable, in addition to the viral load trajectory where the modal Ct value rises above the limit of detection. The probability of being detectable on day $a \leq t_{eclipse} + t_{peak} + t_{switch}$ is $\phi_a = P[C(a) \leq C\_LOD]$ and the probability of being detectable on day $a > t_{eclipse} + t_{peak} + t_{switch}$ is $\phi_a = P[C(a) \leq C_{LOD}](1 - p_{addl})^{a - t_{eclipse} - t_{peak} - t_{switch}}$.

Comprehensive datasets to inform the entire viral load trajectory are currently lacking, and we therefore parameterized the model based on several key features of viral load kinetics that have been determined for SARS-CoV-2 infection. Viral loads and the timing of key events are reported with different reference points (for example, relative to the timing of symptom onset or relative to timing of exposure). However, most existing viral load time series begin after symptom onset (and are therefore from symptomatic individuals), and it is therefore difficult to corroborate assumptions for the pre-symptomatic period and asymptomatic viral trajectory. Fig. S1 depicts where in the disease course these events and parameters may be reported, which we use as a basis to choose parameter values and priors for our model. We discuss the rationale behind this parameterization and the chosen parameter values below. We note that in all analyses, we used informative priors for these key features rather than fixing point estimates, incorporating uncertainty into our inference.

*a) Time from infection to first detectable viral load*

Human infection times are never observed directly, which makes inference of viral kinetics immediately following infection challenging. Data from rhesus macaque challenge and hamster transmission models suggest that viral loads are detectable on the day of infection (*46–48*). In a cat transmission model, some secondary infected cats had detectable viral loads in oropharyngeal secretions on day two post infection (*49*). These studies suggest a very short or non-existent undetectable phase, however, animal models are likely inoculated with much higher viral loads than natural human infection, which may accelerate the time to detection.

We parameterized our viral kinetics model such that 50% of individuals had measured Ct values below the limit of detection at around day two post infection. We assumed that Ct values at the time of infection were distributed around a modal Ct value of 40 and declined thereafter, which resulted in some individuals having detectable viral loads up to five days prior to the time of typical symptom onset, which fits with studies where Ct values have been detected as early as six days pre symptom onset (*50*). For the SEEIRR compartmental model described above, we assumed a mean duration of pre-detectability (*$1/\sigma$*) of two days.

*b) Time from infection to peak viral load*

Challenge studies in rhesus macaques indicate that viral load peaks around two days after infection (*46, 49*). This is not compatible with observations from human data, which suggest that viral loads peak around the time of symptom onset, which typically occurs around five days post infection (see below). Therefore, we assumed that the Ct value reaches a minimum (log viral load peaks) on average five days post infection.

*c) Incubation period for symptom onset*

The time from infection to onset of symptoms has a median incubation period of 5-6 days with 99% of onsets occurring within 14 days (*51*). Although we do not explicitly model symptomatic vs. asymptomatic individuals, this parameter is useful for quantifying other key events where data are usually reported with respect to symptom onset rather than infection. Furthermore, a model comparison analysis by Ferretti et al. found that infectiousness may be tied to the timing of symptom onset rather than time since infection, which supports the parameterization of the viral kinetics curve with respect to time of symptom onset (*51, 52*).

*d) Duration of growth phase and onset of infectiousness*

We assumed that mean Ct decreased monotonically from the time of infection until a minimum at day five post infection, ignoring any eclipse phase ($t_{eclipse} = 0$) before the onset of viral growth. We note that the model may be parameterized such that $t_{eclipse} > 0$, but we simplified this part of the kinetics curve due to limited data and to minimize model complexity. For the SEEIRR model described above, we assumed that individuals took two days on average to transition from the pre-infectious ($E_2$) to the infectious ($I$) compartment ($1/a$). This was chosen such that infected individuals were pre-infectious for four days on average ($1/\sigma + 1/a$), earlier than the time of symptom onset, capturing the observation that a substantial proportion of transmission occurs pre-symptomatically (*15, 50, 53*). For the simpler SEIR model described above, we assumed that individuals were pre-infectious ($E$) for a mean of four days.

*e) Time from peak viral load to symptom onset*

Modelling analyses by He et al. and Ferretti et al. place the most likely time of transmission at around the time of symptom onset, suggesting that infectious viral load and symptom onset may coincide (*51, 53*). Because most viral load data are reported after symptom onset, there is limited pre-symptomatic data to assess if viral titers peak before onset (*54*). However, viral loads and PCR sensitivity appear to decrease monotonically from the time of symptom onset, suggesting that viral loads are highest just after or before the time of symptom onset (*42, 55*). We therefore parameterized our model to place peak viral load at day five post infection, coinciding with the median symptomatic incubation period.

*f) Time from peak viral load to loss of infectiousness*

The vast majority of studies report viral loads, culture-viable virus and model-inferred infectiousness with respect to symptom onset, which may occur after peak viral loads. We therefore quantify the timing of infectiousness with respect to symptom onset, described below.

*g) Time from peak viral load to loss of detectable viral load*

As in f), we quantify waning rates with respect to symptom onset as described below due to the lack of viral load data reported with respect to peak viral load.

*h) Time from symptom onset to loss of infectiousness*

The relationship between viral load and infectiousness are currently unknown for SARS-CoV-2. However, there are currently two main proxies used to estimate time-varying infectiousness: model-based results using generation intervals of known infector-infectee pairs (*51, 53*), and the ability to culture live virus from swab samples taken each day post symptom onset (*51, 56*). Model-based analyses have estimated substantial pre-symptomatic transmission probability, suggesting that individuals are infectious before symptom onset (*51, 53*). For virus culture data, a systematic review found that viable virus is unlikely to be cultured from samples taken more than nine days post symptom onset and another study found that higher viral loads are correlated with probability of live virus culture, though we note that lack of viral culture has not been shown to indicate lack of infectiousness (*15, 54, 57–59*).

For the SEIR transmission model, a rapid scoping review suggested using a median six-day infectious period for asymptomatic infections and median 9.5 days for symptomatic infections (*15, 53*). However, because much of the data we analyze here were collected from populations under transmission-reducing interventions where the infectious period would likely be shorter, we assumed that the observed mean $(1/\gamma)$ infectious period was four days in both the SEIR and SEEIRR models. We note that we do not fix this parameter but estimate it alongside other parameters using a strong prior, and therefore do not exclude the possibility of longer or shorter infectious periods.

*i) Time from symptom onset to loss of detectable viral load*

Waning of viral loads occurs following the onset of symptoms, with the median time from onset to loss of detectability in upper respiratory tract samples of approximately two weeks, though some studies suggest a more prolonged waning rate and greater persistence in lower respiratory tract and sputum samples (*15, 37, 44, 55, 60*). One patient has been reported detectable at day 83 post symptom onset, indicating that some individuals remain PCR positive long after symptom onset (*61*). We therefore parameterized our model to capture both a median time from symptom onset to detectability loss of around two weeks and the possibility for some individuals remaining detectable for much longer. In the SEEIRR model, we used a point estimate for $1/\omega$ (recovery period) of 11 days, corresponding to an average loss of detectability at 19 days post infection (14 days post typical symptom onset).

*Fitting the viral kinetics model*

The literature summarized above informed parameters for the pre-viral peak phase and provided a basis with which to fully parameterize the model. To more formally parameterize the viral kinetics model, we used a least-squares optimization framework to obtain parameter point estimates that gave rise to viral kinetics with the following constraints: the proportion of individuals that are detectable on each day post symptom onset declines in line with existing data (*55*); the lower 99$^{th}$ percentile of possible Ct values at peak viral load is in line with either the lowest observed Ct value in our Brigham & Women's Hospital dataset or from the nursing home data; and the highest 99th percentile of Ct values 30 days post infection is in line with either a Ct value of 32 in the BWH analyses or 30 in the nursing home analyses. We obtained estimates for mean peak viral load in line with the range reported in other studies (up to $10^9$ viral RNA copies per ml) (*43, 62*). We used these point estimates to derive informative priors on key model parameters, as described in Tbl. S1. The resulting distribution of Ct values and detectable proportions at each day $a$ after infection are shown in Fig. S2. These parameters are used as fixed values in Fig. 1.

We note that for the pre-peak phase, solving the model from an initial Ct value at the LOD on day 0 allows for a proportion of individuals to have detectable viral loads up to five days prior to peak viral load (the typical time of symptom onset), which captures individuals who may have longer incubation periods and therefore detectable viral loads for many days before onset.

Single Cross-Section Model

*Likelihood of Daily Probability of Infection*

For a single testing day $t$, let $\pi_{t-A_{max}}, \dots, \pi_{t-1}$ be the marginal daily probabilities of infection for the whole population for $A_{max}$ days to 1 day prior to $t$, respectively, where $t - A_{max}$ is the earliest day of infection that would result in detectable PCR values on the testing day. That is, $\pi_{t-a}$ is the probability that a randomly-selected individual in the population was infected on day $t - a$. Let $p_a(x)$ be the probability that the Ct value is $x$ for a test conducted $a$ days after infection, as described in the Ct model above. As above, let $\phi_a$ be the probability of a Ct value being detectable $a$ days after infection. Let the PCR test results from a sample of $n$ individuals be recorded as $X_1, \dots, X_n$. Then, for $x_i \leq C_{LOD}$ (i.e., a detectable Ct value), the probability of individual $i$ having Ct value $x_i$ is given by:

$$P\left(X_i = x_i \mid \pi_{t-A_{max}}, \dots, \pi_{t-1}\right) = \sum_{a=1}^{A_{max}} p_a(x_i)\pi_{t-a}.$$

The probability of a randomly-chosen individual being detectable to PCR on testing day $t$ is:

$$P\left(X_i \leq C_{LOD} \mid \pi_{t-A_{max}}, \dots, \pi_{t-1}\right) = \sum_{a=1}^{A_{max}} \phi_a \pi_{t-a}.$$

So the likelihood for the $n$ PCR values is given by:

$$\mathcal{L}\left(\pi_{t-A_{max}}, \dots, \pi_{t-a} \mid X_1, \dots, X_n\right) = \prod_{i=1}^{n} \left[ \left( \sum_{a=1}^{A_{max}} p_a(X_i) \pi_{t-a} \right)^{I(X_i \leq C_{LOD})} \left( 1 - \sum_{a=1}^{A_{max}} \phi_a \pi_{t-a} \right)^{I(X_i > C_{LOD})} \right],$$

where $I(\cdot)$ equals 1 if the interior statement is true and 0 if it is false.

If only detectable Ct values are recorded as $X_1, \dots, X_n$, then the likelihood function is given by:

$$\mathcal{L}\left(\pi_{t-A_{max}}, \dots, \pi_{t-1} \mid X_1, \dots, X_n\right) = \prod_{i=1}^{n} \left[ \frac{\sum_{a=1}^{A_{max}} p_a(X_i) \pi_{t-a}}{\sum_{a=1}^{A_{max}} \phi_a \pi_{t-a}} \right] = \frac{\prod_{i=1}^{n} \left[ \sum_{a=1}^{A_{max}} p_a(X_i) \pi_{t-a} \right]}{\left( \sum_{a=1}^{A_{max}} \phi_a \pi_{t-a} \right)^n}.$$

Either of these likelihoods can be maximized to get nonparametric estimates of the daily probability of infection, with the constraint that $\sum_{a=1}^{A_{max}} \pi_{t-a} \leq 1$. To improve power and interpretability of the estimates, however, we consider two parametric models.

*Exponential Growth Model*

Assume that over the $A_{max}$ days prior to testing day *t*, the daily incidence grows (or declines) exponentially at rate $\beta$ so that $\pi_{t-a} = \pi_0 \exp[\beta(t-a)]$. The exponential growth rate $\beta$ is thus the logarithm of the daily growth rate in incidence over the days $t - A_{max}$ to $t-1$. Larger values of $\beta$ indicate faster growth of new infections and a value of 0 indicates no increase or decrease in the number of new infections each day. For positive values of $\beta$, the doubling time (in days) for new infections is given by $\frac{\log 2}{\beta}$. This model may be a reasonable approximation in the early stages of an outbreak, when the number of susceptible individuals is large compared to the number of infections (*19*).

Fitting this likelihood, and ignoring the nuisance parameter $\pi_0$, gives:

$$\mathcal{L}(\beta \mid X_1, \dots, X_n) \propto \prod_{i=1}^{n} \left[ \left( \sum_{a=1}^{A_{max}} p_a(X_i) \exp[\beta(t-a)] \right)^{I(X_i \leq C_{LOD})} \left( 1 - \sum_{a=1}^{A_{max}} \phi_a \exp[\beta(t-a)] \right)^{I(X_i > C_{LOD})} \right],$$

if detectable and undetectable Ct values are recorded. If only detectable Ct values are recorded, then the parametric likelihood is given by:

$$\mathcal{L}(\beta \mid X_1, \dots, X_n) \propto \frac{\prod_{i=1}^{n} \left[ \sum_{a=1}^{A_{max}} p_a(X_i) \exp[\beta(t-a)] \right]}{\left( \sum_{a=1}^{A_{max}} \phi_a \exp[\beta(t-a)] \right)^n}.$$

To incorporate uncertainty in the distribution of viral loads on each day after infection, we construct a Bayesian framework for estimation and inference. In this paper, we use a normally-distributed prior for $\beta$ with mean 0 and standard deviation 0.25, fitted using a Markov chain Monte Carlo (MCMC) algorithm to obtain the posterior distribution. Note that $\beta = 0.1$ corresponds to a doubling time for new infections of approximately one week. The prior distributions for the parameters for the Ct kinetics model are given in Tbl. S1 and described above.

This method also results in posterior distributions for the parameters for the Ct kinetics model. These are nuisance parameters for estimation of the epidemic trajectory but may be useful in improving the priors for future use of this model.

*SEIR Model*

As an alternative parameterization of the likelihood, we also use the SEIR compartmental model described above (see *Simulated Epidemic Transmission Model*). We generate Ct values for all exposed, infectious, and recovered individuals when they are sampled based on the Ct value model described above. This model is most appropriate for a relatively closed population, where the outbreak is initiated by one or several initial infections and no transmission-reducing measures are taken over the time period studied. We use this model for the nursing home outbreaks observed early in the SARS-CoV-2 pandemic in Massachusetts and the single cross-section analyses using the BWH data.

The prior distributions for the parameters of the SEIR model are given in Tbl. S1. For any set of parameters drawn from these prior distributions, the probability of infection is determined, and the likelihood found using the appropriate nonparametric likelihood equation (either including all samples or only samples with a detectable Ct value) given above. Posterior distributions for the SEIR model parameters are obtained using MCMC fitting, along with posterior distributions for the Ct kinetics model parameters. From the posterior estimates of the SEIR model parameters, posterior distributions can also be found for related model features, such as $R_t$, as of the testing day $t$ (*19, 28*).

Simulated Nursing Home Outbreaks

To ensure that our method provides accurate estimates of the epidemic trajectory, we performed extensive simulation-recovery experiments using a synthetic nursing home population undergoing a stochastic SEIR epidemic (Fig. S6A). We also extended our method to use only the distribution of detectable Ct values, providing an alternative approach where only the results from positive samples are recorded. We then evaluated the accuracy and precision of growth rate estimates from the SEIR and exponential growth models fitting to cross-sectional Ct values observed during epidemic growth, decline and around the peak (Fig. S6B–D). The SEIR model consistently provided unbiased, constrained daily growth rate estimates at all three timepoints when all sample results were used. When only the distribution of detectable Ct values were used, estimates during the growth and decline phases were accurate but exhibited wide credible intervals, whereas estimates during the peak phase were slightly biased upward. Estimates of the average growth rate from the exponential model were consistent using either all or only positive samples—slightly higher than the daily growth rates from the SEIR model during the peak and decline phases, reflecting the drop in daily growth rate relative to the path average as the epidemic begins to decline.

We also found that increasing the simulated population size from 300 to 5000 did not change the accuracy of our estimates and had only a modest impact on 95% credible interval widths (Fig. S7). Similarly, using progressively less informative priors for the viral kinetics parameters did not change the accuracy of the inferred growth rates, but did increase the uncertainty in the growth rate estimates (Fig. S8).

Multiple Cross-Sections Model

Now we consider settings where there are multiple days of testing, $t_1, \ldots, t_T$. We again denote by $\pi_t$ the probability of infection on day $t$ and now denote the sampled Ct value for the $i^{\text{th}}$ individual sampled on test day $t_j$ by $X_i^{t_j}$, where $i \in 1, \ldots, n_j$ for test day $j$ and $j \in 1, \ldots, T$. Note that individual $i$ may refer to different individuals on different testing days. Let $\{\pi_t\}$ be the daily probabilities of infection for any day $t$ where an infection on day $t$ could be detectable using a PCR test on one of the testing days. By a straightforward extension of the likelihood for the single cross-section model, the nonparametric likelihood for the set of infection probabilities $\{\pi_t\}$, when samples with and without a detectable Ct value are included, is given by:

$$\mathcal{L}(\{\pi_t\} \mid X_1^{t_1}, \ldots, X_{n_1}^{t_1}, \ldots, X_{n_T}^{t_T}) = \prod_{j=1}^{T} \left\{ \prod_{i=1}^{n_j} \left[ \left( \sum_{a=1}^{A_{max}} p_a\left(X_i^{t_j}\right) \pi_{t_j-a} \right)^{I\left(X_i^{t_j} \leq C_{LOD}\right)} \left( 1 - \sum_{a=1}^{A_{max}} \phi_a \pi_{t_j-a} \right)^{I\left(X_i^{t_j} > C_{LOD}\right)} \right] \right\}$$
$$= \prod_{j=1}^{T} \left\{ \left[ \prod_{i=1}^{n_j} \left( \sum_{a=1}^{A_{max}} p_a\left(X_i^{t_j}\right) \pi_{t_j-a} \right)^{I\left(X_i^{t_j} \leq C_{LOD}\right)} \right] \left[ 1 - \sum_{a=1}^{A_{max}} \phi_a \pi_{t_j-a} \right]^{n_j^-} \right\},$$

where $n_j^-$ is the number of undetectable samples on testing day $t_j$.

Only considering samples with a detectable Ct value gives the likelihood:

$$\mathcal{L}(\{\pi_t\} \mid X_1^{t_1}, \ldots, X_{n_1}^{t_1}, \ldots, X_{n_T}^{t_T}) = \prod_{j=1}^{T} \left\{ \frac{\prod_{i=1}^{n_j} \left[ \sum_{a=1}^{A_{max}} p_a\left(X_i^{t_j}\right) \pi_{t_j-a} \right]}{\left[ \sum_{a=1}^{A_{max}} \phi_a \pi_{t_j-a} \right]^{n_j}} \right\}.$$

Either of these likelihoods can be parameterized using the exponential growth rate model described above. However, the exponential growth rate model is less likely to be a good approximation of the true incidence probabilities over a longer period of time, so it may not be a good model for multiple test days that cover a long stretch of time.

The SEIR model can be used with multiple testing days as well. It is fit as described above with one of these likelihoods in place of the single cross-section model likelihood, with posterior distribution estimates obtained via MCMC fitting.

Markov Chain Monte Carlo Framework

We used an MCMC framework with the Metropolis-Hastings algorithm to generate either multivariate Gaussian or univariate uniform proposals. For all single-cross section analyses (Figs. 2,3), we used a modified version of this framework with parallel tempering: an extension of the algorithm that uses multiple parallel chains to improve sampling of multimodal posterior distributions (*63*). For the multiple cross section analyses in Fig. 4, we used the unmodified Metropolis-Hastings algorithm because the computational time of the parallel tempering algorithm is far longer, and these analyses were underpinned by more data and less affected by multimodality. In all analyses, three chains were run upward of 80,000 iterations (500,000 iterations for the Gaussian process models). Convergence was assessed based on all estimated parameters having an effective sample size greater than 200 and a potential scale reduction factor ($\hat{R}$) of less than 1.1, evaluated using the *coda* R package (*64*).

Simulated Testing Schemes

Standard approaches to estimating doubling time, growth rate, or $R_t$ are subject to misestimation as a result of changes in testing policies (*5*). To assess the effect of such changes on our methods, we simulate changes in testing rates in Fig. 3 and assess the effect on $R_t$ estimation using *EpiNow2* with reported case counts and using Ct-based methods with surveillance samples.

The reported case counts arise from a model of symptom onset and reporting delays. We assume a log-normal incubation period with mean of log(5) days and standard deviation 0.418, as estimated by Lauer et al. (*52*). Each symptomatic individual has some probability of being tested after a reporting delay, where this probability may vary by day of the outbreak. Three scenarios are considered: flat testing (fixed probability of testing of 10%); increasing testing rates (a linear increase in probability of testing from 10% 36 days prior to the analysis day to 20% one day prior to the analysis day); decreasing testing rates (a linear decrease in probability of testing from 10% 36 days prior to the analysis day to 1% one day prior to the analysis day).

$R_t$ is estimated from these simulated data using the R package *EpiNow2* (*28, 31*, https://github.com/epiforecasts/EpiNow2). This requires the following inputs:

1. Time series data for the number of newly confirmed cases per day.
2. A specified incubation period distribution, giving the distribution of delays between infection and symptom onset.
3. A specified reporting delay distribution, giving the distribution of delays between symptom onset and case confirmation.

4. Priors on the generation interval distribution, specifying the mean and standard deviation of the times between infection in infector-infectee pairs.

For the reporting delay distribution, we assumed a discretized gamma distribution with shape and scale parameters of 5 and 2 respectively (mean of 2.5 days and standard deviation of 1.12 days). For the generation interval, in our simulations using the SEIR model, the mean generation interval is given by $T_c = 1/\sigma + 1/\gamma$, where $\sigma$ and $\gamma$ are the inverse of the mean incubation and infectious periods in days, respectively. The variance of the generation interval distribution is given by $Var = 2\left(\frac{T_c}{2}\right)^2$. Therefore, $T_c$ =8 days, with standard deviation of 5.66 days. In *EpiNow2*, normal priors were placed on these quantities with standard deviations of 3 for both.

For the surveillance Ct sample analyses, we use random sampling where each individual has a 0.3% probability of being tested at some point in the outbreak. We consider a single testing day (sampling 0.3% of the population), two testing days one week apart (sampling 0.15% of the population on each day), and three testing days each one week apart. For the three testing days, we consider scenarios where the probability of being sampled on any one of those days is flat at approximately 0.1%, rising from 0.05% to 0.10% to 0.15%, or falling from 0.15% to 0.10% to 0.05% across the three days. In all of these settings, approximately 3000 total tests are conducted. Fig. 3B plots the estimates using these methods from 100 simulations at two time points in the epidemic: one before the peak incidence (true $R_t > 1$) and one after the peak incidence (true $R_t < 1$).

Epidemic Seed Time Priors

External information on the epidemiological context can be used to further constrain estimated epidemic trajectories. When fitting the SEIR model to the single cross-sections of Ct values in Massachusetts, we are estimating the dynamics of a single epidemic peak that precedes the observation time. We therefore placed uniform priors on the epidemic seed time, $t_0$, to reflect prior knowledge of the start of the two epidemic growth phases. Specifically, for samples taken prior to 2020-06-01, we assumed that the seed time was between 2020-02-01 and 2020-03-01. For samples taken between 2020-06-01 and 2020-08-01, we assume that the seed time is unknown between 2020-02-01 and two weeks prior to the sample time. This captures the assumption that we do not know if infections during this time are dominated by the decline phase of the first wave or the growth phase of the second. If the sample time was after 2020-08-01, we assume that the seed time of the second wave is unknown between 2020-06-01 and 2 weeks prior to the sample time. This ensures that we are estimating incidence based on the second wave for these later samples.

As a sensitivity analysis, we instead assumed that the epidemic seed time was unknown between 2020-02-01 and the sampling date for all sampling times (Fig. S10D,E). Estimated growth rates were very similar, though the posterior densities for the first four weeks were wider when the epidemic seed times were less constrained. Some of the posterior estimates were bimodal (Fig. S11), where the same Ct distribution could be explained as resulting from very recent and fast epidemic growth (most high Ct values are from the upswing of a recent infection) or from the downswing of a declining epidemic (most high Ct values are from the clearance phase). Although our method is able to accurately estimate these bimodal posterior distributions, it is important to interpret them alongside the epidemiological context. Without suitable priors, estimates that are mathematically correct are not necessarily epidemiologically plausible.

Gaussian Process Model

For a highly general version of our method, we use a Gaussian Process (GP) prior for the daily incidence rate, which is denoted $\beta(t)$, for any day $t$ where infection on that day could result in a positive PCR test on the (or one of the) testing day(s). For identifiability, the sum of $\beta(t)$ over all possible values of $t$ is set equal to 1; thus, the resulting estimates should be considered the relative probability of infection on each day, relative to the set of possible days. When only positive PCR test results are included in the inference, we estimate $\beta(t)$ directly. When negative PCR tests are also included, we multiply $\beta(t)$ by an estimated scaling factor between 0 and 1, which is the absolute probability of infection from the entire incidence curve. Xu et al. describe and illustrate the use of the GP as a prior distribution for nonparametric inference on incidence rates for various infectious disease settings (*32*). We use the GP prior for the vector of values $k(t)$ for each day $t$; the incidence rate for day $t$ is then given by $\beta(t) = \left(1 + e^{-k(t)}\right)^{-1}$. The covariance matrix for the GP prior $k \sim MVN\left((0, \dots, 0), \boldsymbol{K}\right)$ is given by $\boldsymbol{K}_{ij} = \eta^2 \exp\left(-\rho^2 D_{ij}^2\right)$, where $D_{ij}$ is the difference between $i$ and $j$ and $\nu$ and $\rho$ are hyperparameters with exponentially-distributed priors with means of 1.5 and 0.03, respectively (see Tbl. S1). The parameter $\rho$ determines the rate of decline of the covariance as the time between days increases, so a higher value of $\rho$ indicates less correlation. See McElreath for more details (*65*). We present results from this model in terms of the posterior distribution for the $\beta(t)$, the daily incidence rate values.

This method allows for more flexibility than the one based on the SEIR model, allowing the daily incidence rate to reflect changes in transmission rates and contact patterns (although these are not separately identified by the model), as well as the depletion of the pool of susceptible individuals. Thus it does not require the strict parametric assumptions of the SEIR model. We use it to model the course of the outbreak in Massachusetts, as various policies and behavior changes affect the trajectory over time.

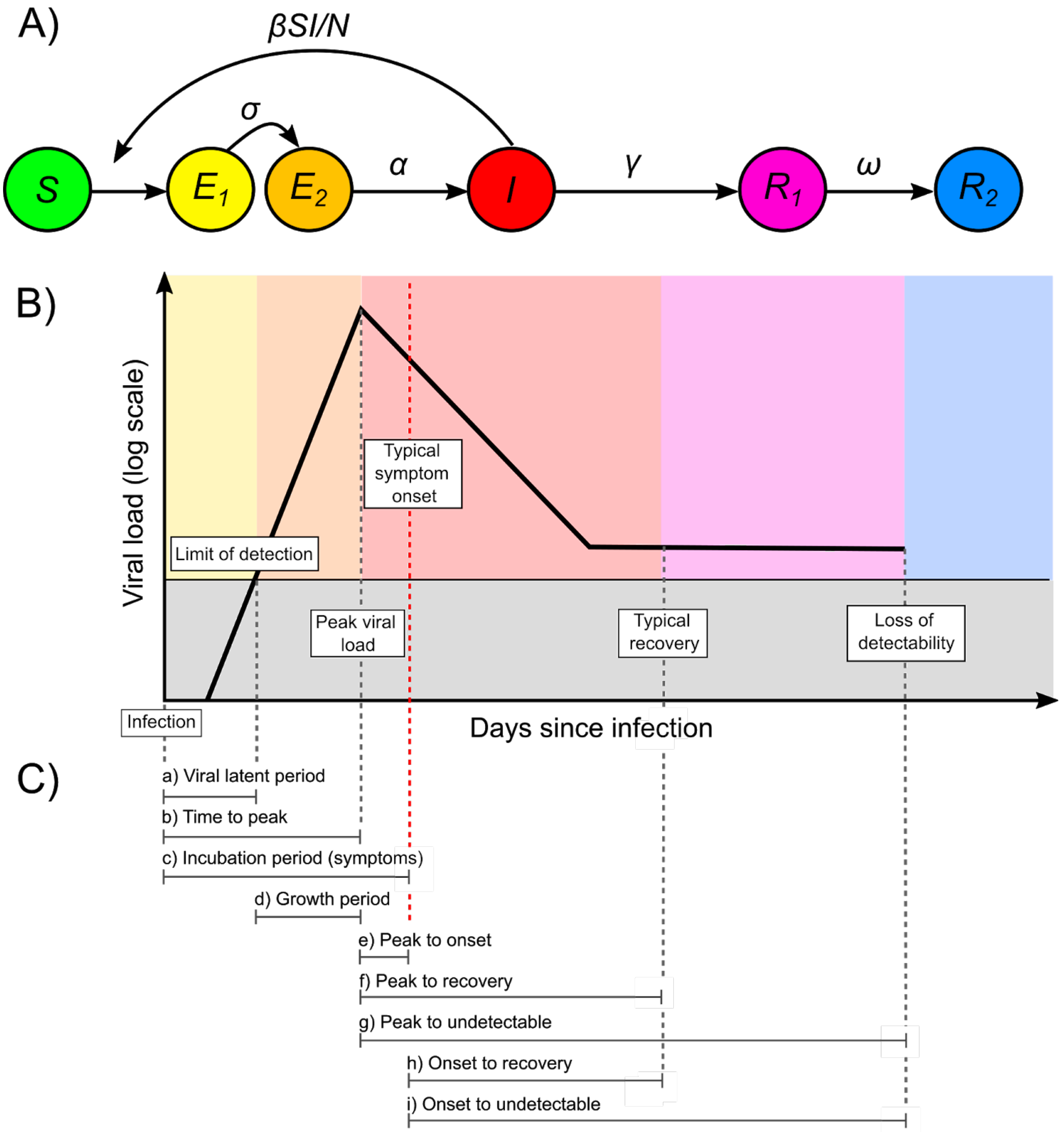


**Fig. S1. Schematic of the SEEIRR transmission model, viral kinetics model and periods of viral kinetics used to parameterize the viral kinetics model.** (**A**) Transmission model with the following compartments: susceptible (*S*), exposed, not infectious, undetectable ($E_1$), exposed, not infectious, detectable ($E_2$), infectious and detectable (*I*), recovered still detectable ($R_1$), and recovered undetectable ($R_2$). Transition rates were as follows: $\beta$, the transmission rate, $\sigma$, onset of detectability rate, $\alpha$, the onset of infectiousness rate, $\gamma$, the rate of infectiousness loss, and $\omega$, the rate of loss of detectability after recovery. Note that the simpler SEIR model effectively combined the $E_1$ and $E_2$ compartments and the $R_1$ and $R_2$ compartments. (**B**) Schematic of the assumed viral kinetics model with key event times labeled. (**C**) Possible time between events for which data might be available.

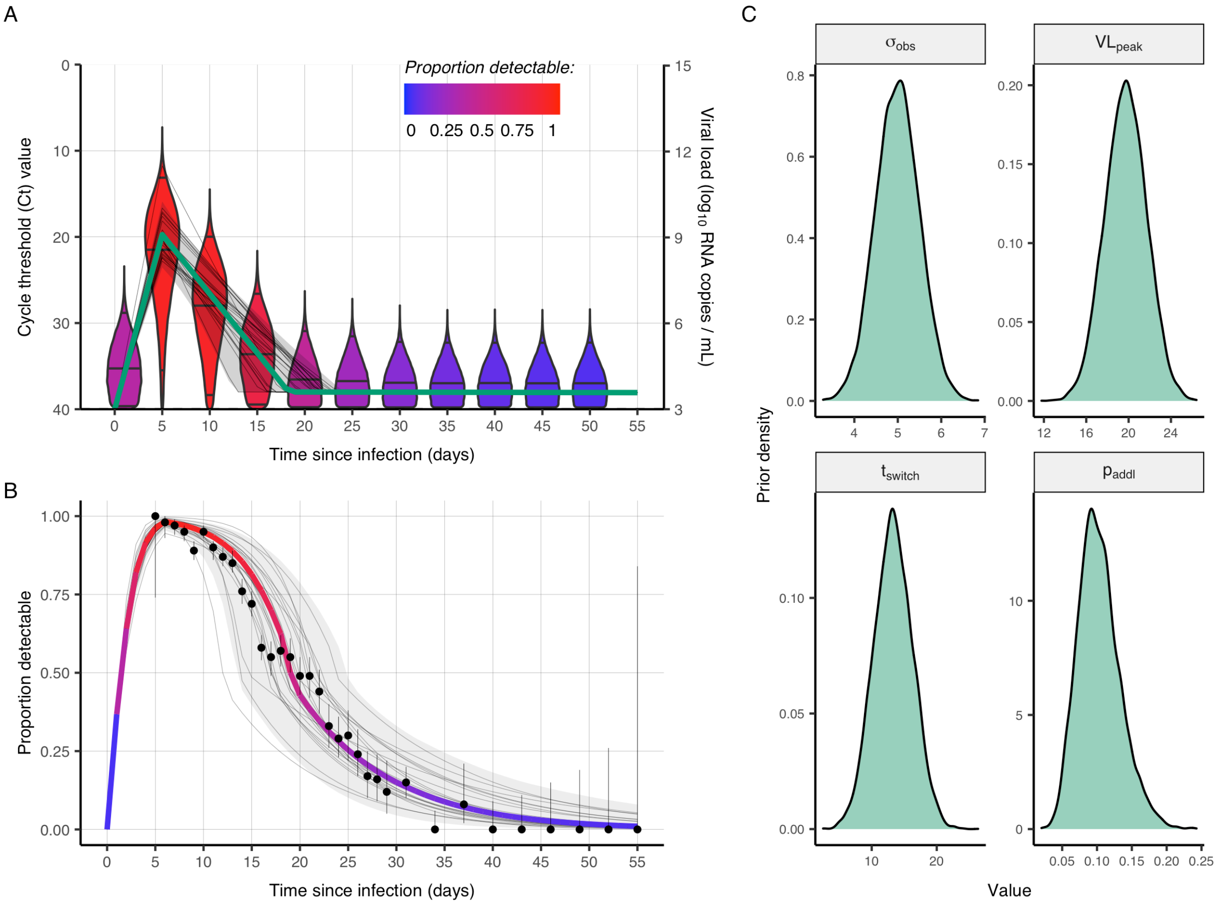


**Fig. S2. Fitted viral kinetics model for cycle threshold (Ct) values, assumed loss of detectability over time and assumed priors on model parameters.** (**A**) Solid green line shows the fitted modal viral load trajectory over time since infection. Faint grey lines show trajectories from prior draws, and faint grey ribbon shows 95% quantiles. Violin plots show the distribution of detectable Ct values for each five-day increment post infection using the maximum *a priori* trajectory (green line). Violins are colored by the proportion of Ct values above the limit of detection. (**B**) Least-squares based fit (colored line) to the proportion of individuals detectable in upper respiratory tract samples on each day post symptom onset (assuming an incubation period of five days) from Borremans et al. (*55*). Black dots and lines show proportion positive and 95% confidence intervals. Faint grey lines show proportion detectable over time from prior draws, and faint grey ribbon shows 95% quantiles. (**C**) Assumed prior densities for unknown model parameters.

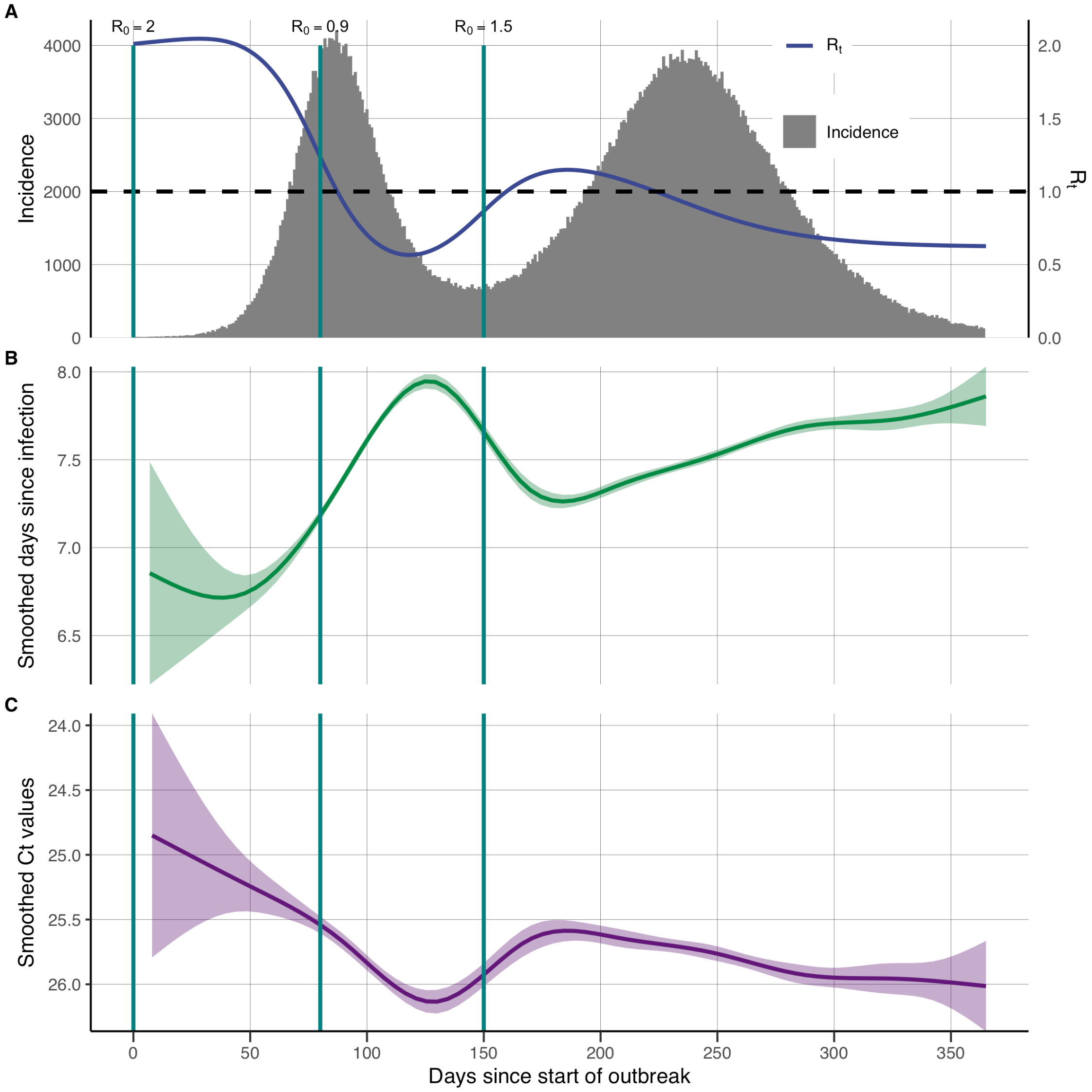


**Fig. S3. The Ct value distribution is expected to change over an epidemic even under symptom-based surveillance.** (**A**) A stochastic SEIR model was simulated with switch points at days 80 and 150, representing changes in $R_0$ driven by implementation of non-pharmaceutical interventions or increasing virus transmissibility. $R_0$ changes were interpolated to smooth transitions between epidemic stages. Teal lines show timing of $R_0$ switch points. (**B**) Symptom-based surveillance was simulated assuming that each infected individual had a 35% change of becoming symptomatic, had an incubation period drawn from a log-normal distribution (*52*), and had a delay between onset and testing drawn from a gamma distribution with mean 2.5 days and variance 1.25 days. The result is a changing distribution of delays between infection and sampling time over the course of the epidemic. The green line and ribbon show LOESS smoothing spline and 95% confidence intervals, respectively, fitted to all simulated delays. (**C**) Each sampled individual has a simulated Ct value based on the time since infection. The purple line and ribbon show LOESS smoothing spline and 95% confidence intervals, respectively, fitted to all simulated Ct values.

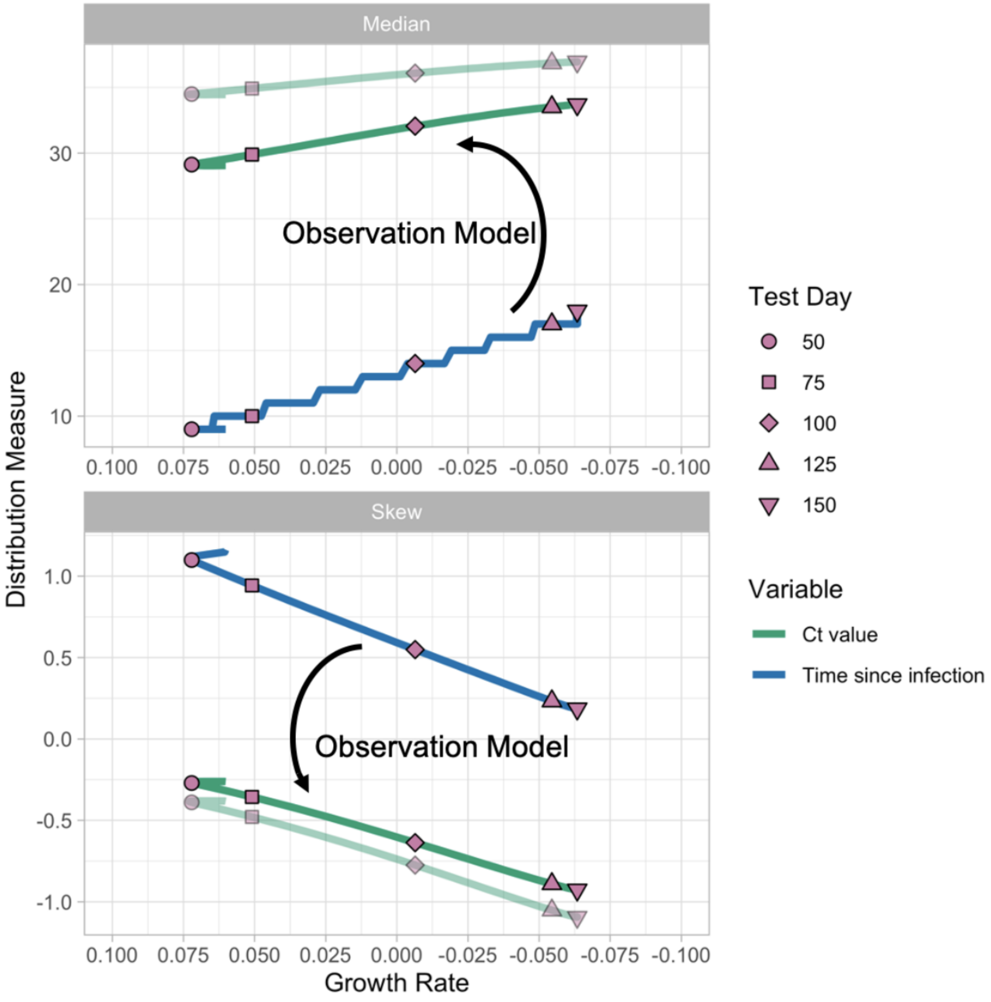


**Fig. S4. Distributional properties of times since infection (blue line) correlate with epidemic growth rate, which can be observed using cycle threshold (Ct) values as a proxy with a properly calibrated observation model (green line).** Median (top) and skewness (bottom) of the time since infection (blue line) and observed Ct value (green lines) by average 35-day growth rate from the simulated susceptible-exposed-infectious-recovered (SEIR) model. The two green lines represent two possible observation models (e.g., from different RT-qPCR machines, protocols, or swab locations).

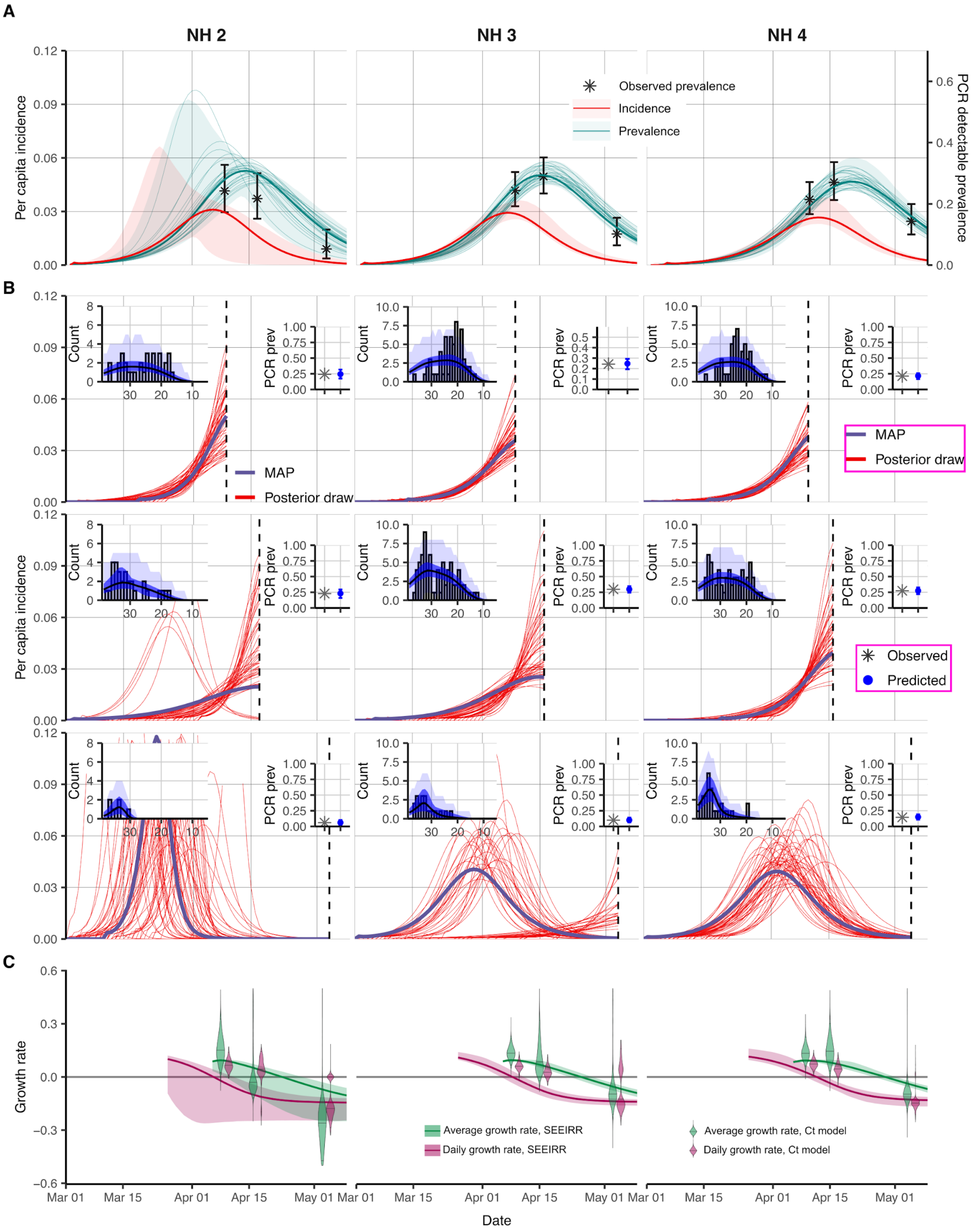


**Fig. S5. Single cross-sectional distributions of observed cycle threshold (Ct) values used to reconstruct epidemic trajectories in three additional Massachusetts nursing homes. (A)** Posterior distribution of prevalence (teal lines and shaded ribbon) from the SEEIRR model fit to point prevalence at three sampling times for each nursing home and posterior distribution of daily per capita incidence (red line and shaded ribbon) from the same model. Black error bars show 95% binomial confidence intervals on PCR positive prevalence. (**B**) Each panel shows results from fitting the Ct-based SEIR model separately to cross-sections of virologic data. Shown are random posterior samples (red lines) and the maximum posterior probability trajectory (purple line) for the incidence curve. Note that we use a parallel tempering algorithm, so conflicting trajectories are an accurate representation of the multi-modal posterior. Left-hand insets show model-predicted Ct distributions (blue) fitted to the observed Ct values (grey bars) from that cross-sectional sample. Posterior median (black line) and 95% CrI for the expected Ct distribution (dark blue ribbon), and 95% prediction intervals based on simulated observations (light blue ribbon). Right-hand insets show model-predicted median (blue point) and 95% CrI (blue error bars) for the proportion of samples testing positive compared to the observed proportion tested positive (grey cross). (**C**) 35-day (green) and 1-day (magenta) average growth rates from Ct value fit at three time points (violin plots) and fit to point prevalence (lines and shaded ribbons) for each nursing home. The three nursing homes included here are different from the one shown in Fig. 2.

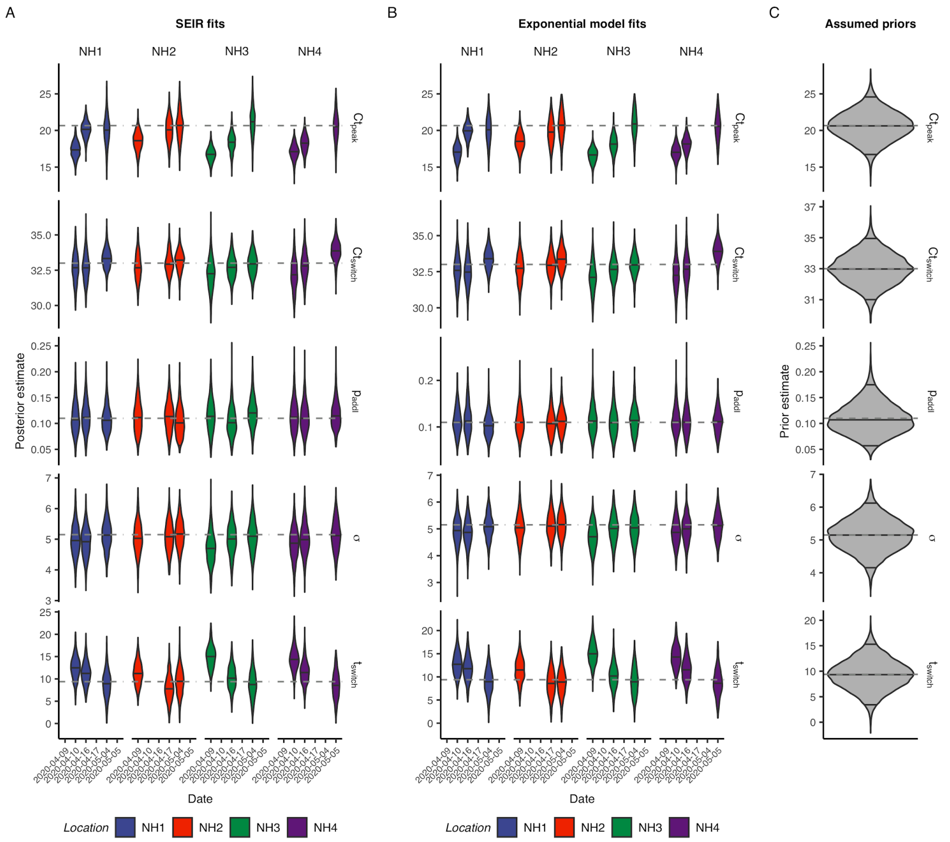


**Fig. S6. Prior and posterior distributions of parameters for models fit to observed cycle threshold (Ct) values from four Massachusetts nursing homes.** (**A** and **B**) Posterior distributions of model parameters from the susceptible-exposed-infectious-recovered (SEIR) model (**A**) and exponential growth model (**B**). (**C**) Prior distributions of model parameters used in both models. NH1 is the nursing home shown in Fig. 2; the others are shown in Fig. S5.

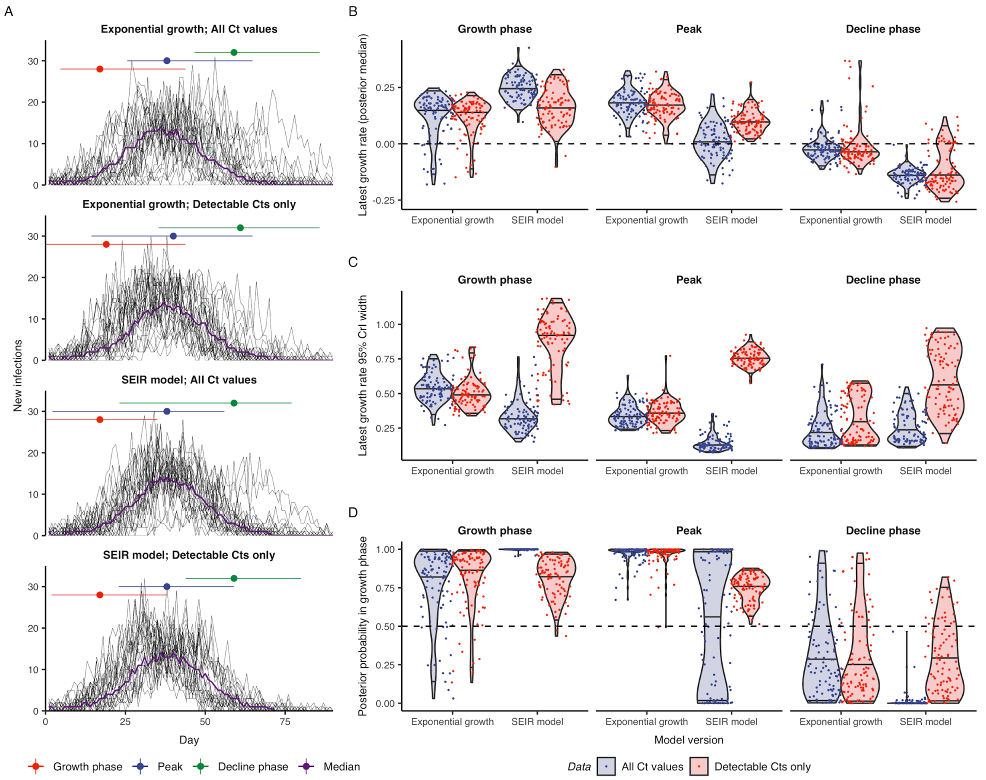


**Fig. S7. In simulations similar to the Massachusetts nursing homes, the cycle threshold (Ct) value-based methods for epidemic trajectory estimation recover the simulation parameters for exponential growth and SEIR models with or without negative test results.** (**A**) Distribution across 100 simulations of median posterior estimates of daily infection incidence by model used with notations of the growth phase, peak, and decline phase of the epidemic. (**B–D**) Distribution across 100 simulations of median posterior estimates of growth rate (**B**), widths of 95% credible intervals of growth rate (**C**), and posterior probability that the growth rate is greater than 0 (**D**) at the end of the three phases of the epidemic for each of four models.

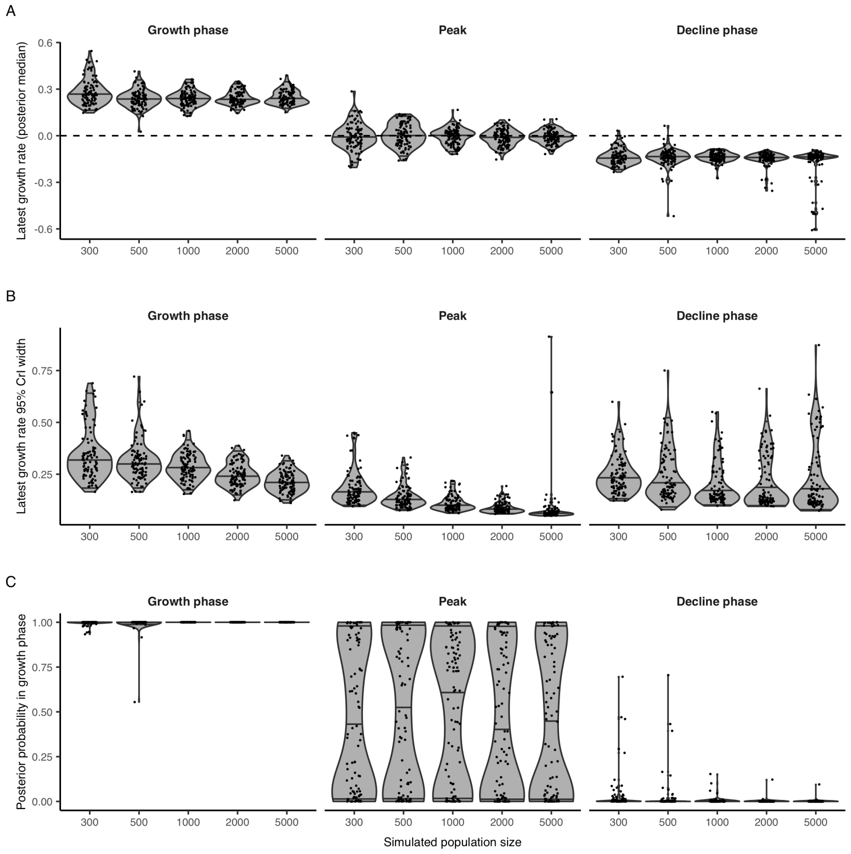


**Fig. S8. Varying population size has little impact on the performance of the cycle threshold (Ct) value-based methods for epidemic trajectory estimation using an SEIR model with negative test results in recovering simulation parameters similar to the Massachusetts nursing home parameters.** Distribution across 100 simulations of median posterior estimates of growth rate (**A**), widths of 95% credible intervals of growth rate (**B**), and posterior probability that the growth rate is greater than 0 (**C**) at the end of the three phases of the epidemic by population size. Results shown are as in Fig. S7B–D, with population size as an additional x-axis.

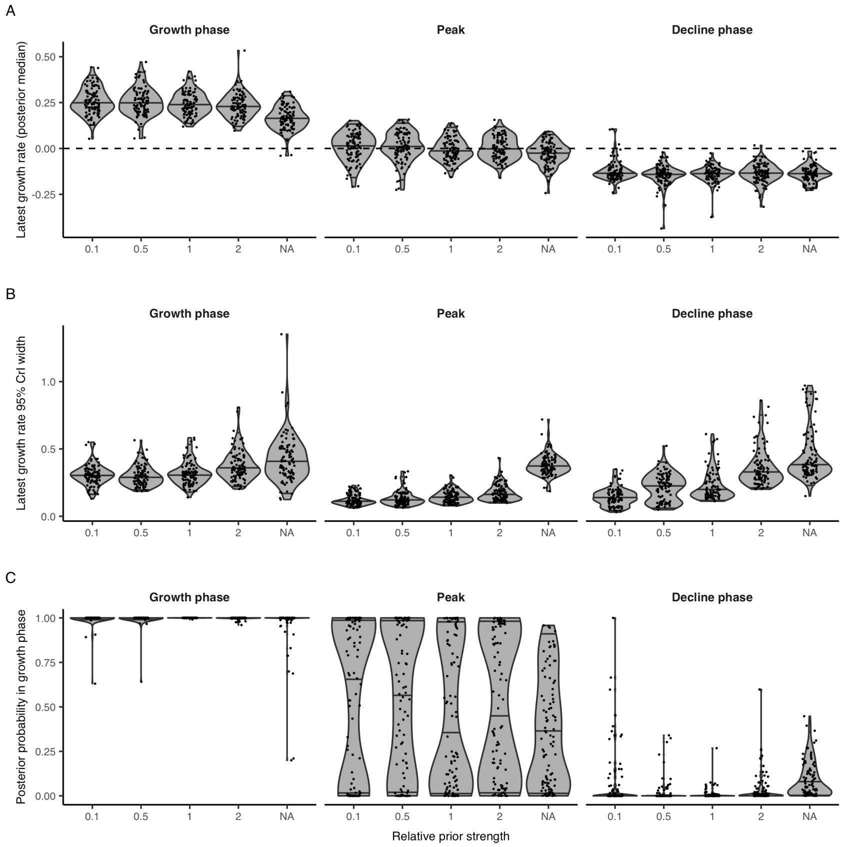


**Fig. S9. Varying the relative strength of the prior distributions has little impact on the performance of the cycle threshold (Ct) value-based methods for epidemic trajectory estimation using an SEIR model with negative test results in recovering simulation parameters similar to the Massachusetts nursing home parameters.** Distribution across 100 simulations of median posterior estimates of growth rate (**A**), widths of 95% credible intervals of growth rate (**B**), and posterior probability that the growth rate is greater than 0 (**C**) at the end of the three phases of the epidemic by relative strength of prior distribution. The rightmost estimates in each plot (labeled NA) indicate the use of uniform priors on all model parameters. Results shown are as in Fig. S7B–D, with relative prior strength as an additional x-axis.

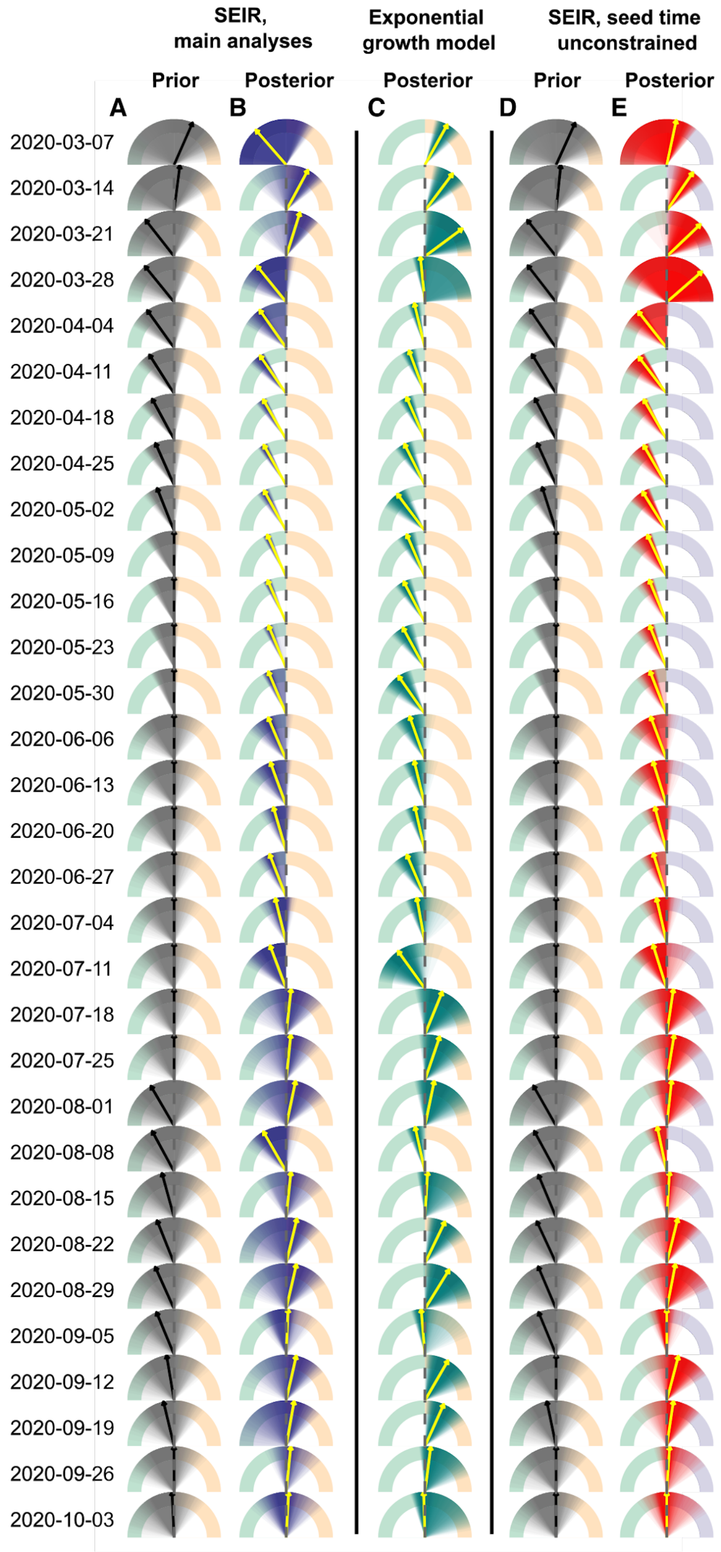


**Fig. S10. All single cross-section growth rate estimates using Ct values from Brigham & Women's Hospital, Massachusetts corresponding to Fig. 4E.** Dials show estimated growth rates using data collected in the week shown to the left of the plot. Dials range from -0.5 to +0.5 (i.e., left of the vertical axis indicates epidemic decline). Shaded regions show posterior or prior densities. Yellow arrows show posterior medians and black arrows show prior medians. Note that wide estimates represent true multimodal posterior distributions, as these Ct distributions could be generated either at the start of fast growth or in decline phases (see Fig. S11). (**A**) Prior daily growth rates (grey) from the SEIR model assuming constraints on the epidemic seed time. (**B**) Posterior daily growth rates (blue) from the SEIR model assuming constraints on the epidemic seed time, as in Fig. 4. (**C**) Posterior 35-day average growth rates (teal) from the exponential growth model. (**D**) Prior daily growth rates (grey) from the SEIR model, assuming no constraints on the epidemic seed time. (**E**) Posterior daily growth rates (red) from the SEIR model, assuming no constraints on the epidemic seed time.

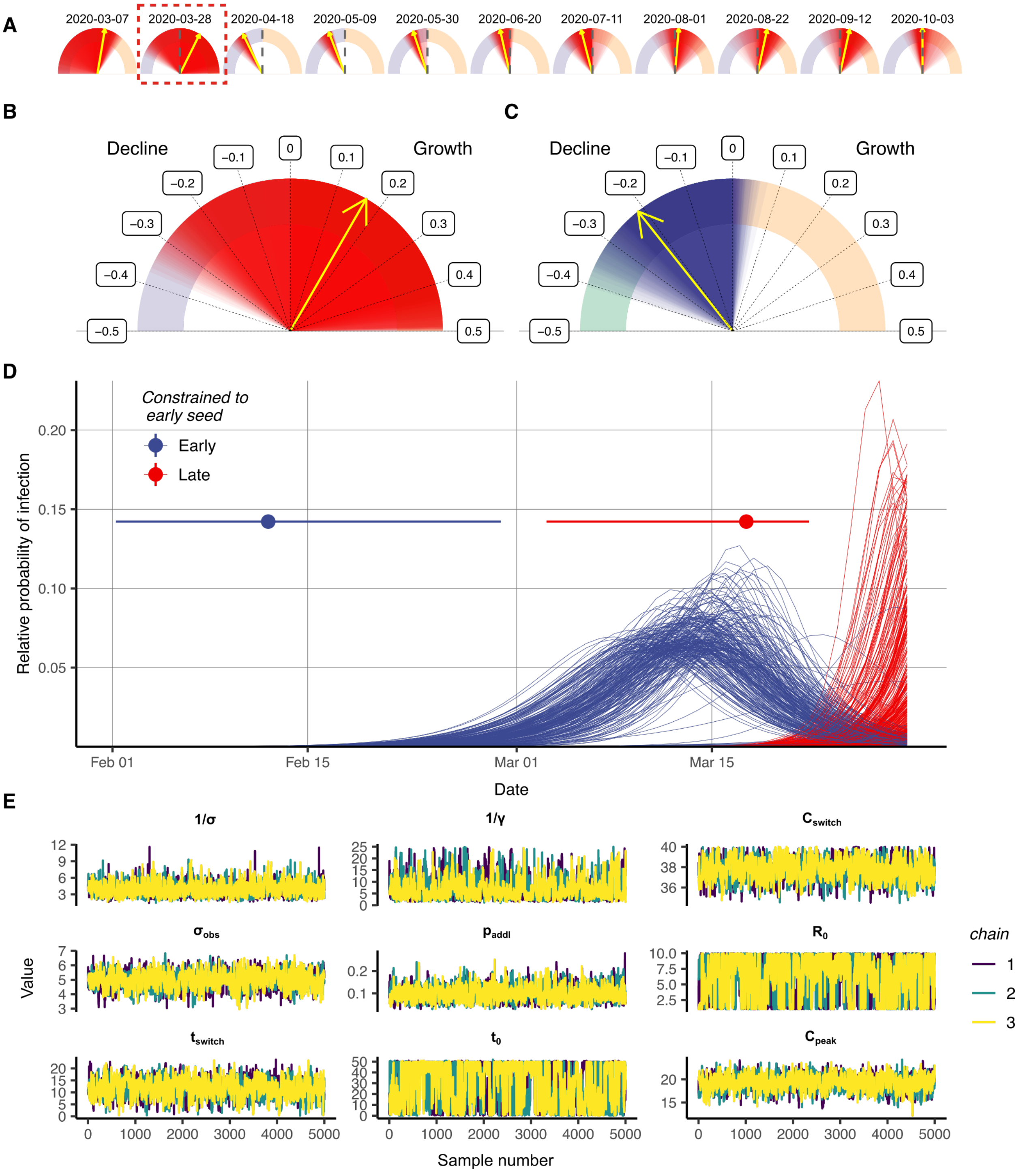


**Fig. S11**. **Estimated epidemic trajectories from single cross sectional Ct distributions can be multimodal.** (**A**) Estimated daily growth rates using single cross-sections of Ct values and the SEIR model with no constraints on the epidemic seed time, matching the same dates shown in Fig. 4E. Red shaded region shows posterior densities, yellow arrows show posterior medians. (**B**) Detailed view of the second dial, 2020-03-28, in (**A**). (**C**) Dial for 2020-03-28 matching Fig. 4E, assuming tighter constraints on the epidemic seed time. (**D**) Epidemic trajectory using BWH data sampled in the week commencing 2020-05-03. Each line is a randomly drawn posterior sample for the SEIR incidence curve using the posteriors shown in (**B**). Lines are colored based on whether the seed time was before 2020-03-01 (Early) or after (Late). Pointrange plots show 95% credible intervals and posterior median on the epidemic seed time depending on whether the seed time was before or after 2020-03-01, demonstrating a multimodal posterior. (**E**) MCMC trace plots for estimated parameters underpinning the trajectories in (**B**). Trace plots demonstrate good convergence, but clear multi-modality for $R_0$ and $t_0$; the same data can be explained with either high $R_0$ and late $t_0$, or low $R_0$ and early $t_0$.

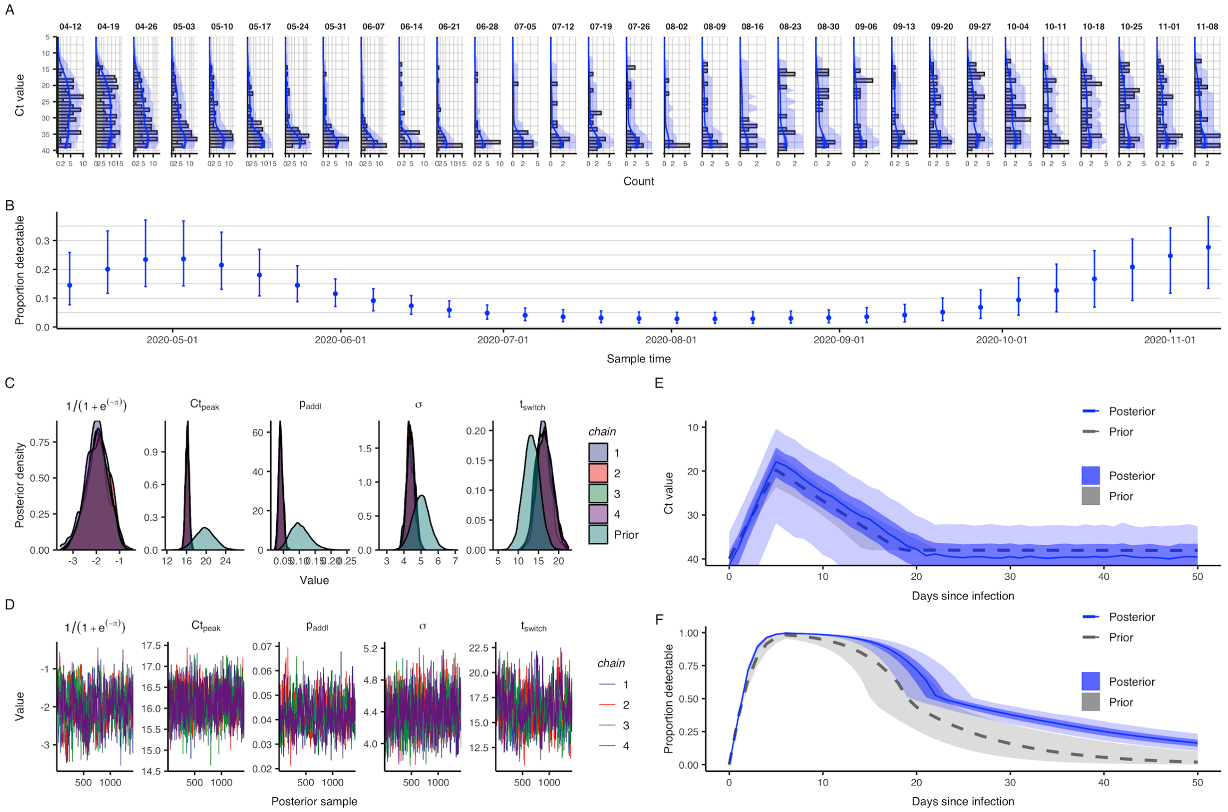


**Fig. S12. Data and results summaries for estimation of epidemic trajectory in Massachusetts shown in Fig. 4.** (**A**) Histogram of observed cycle threshold (Ct) values by weekly sample. (**B**) Proportion of samples with detectable Ct value by weekly sample. (**C**) Prior vs. posterior density of model parameters from Gaussian process model fit to observed Ct value data. (**D**) Markov chain Monte Carlo sampling chains for model parameters fit to observed Ct value data. (**E,F**) Prior vs. posterior density of population-level observed Ct value model, mean Ct value (**E**) and probability of having a detectable Ct value (**F**), by days since infection fit to observed Ct value data.

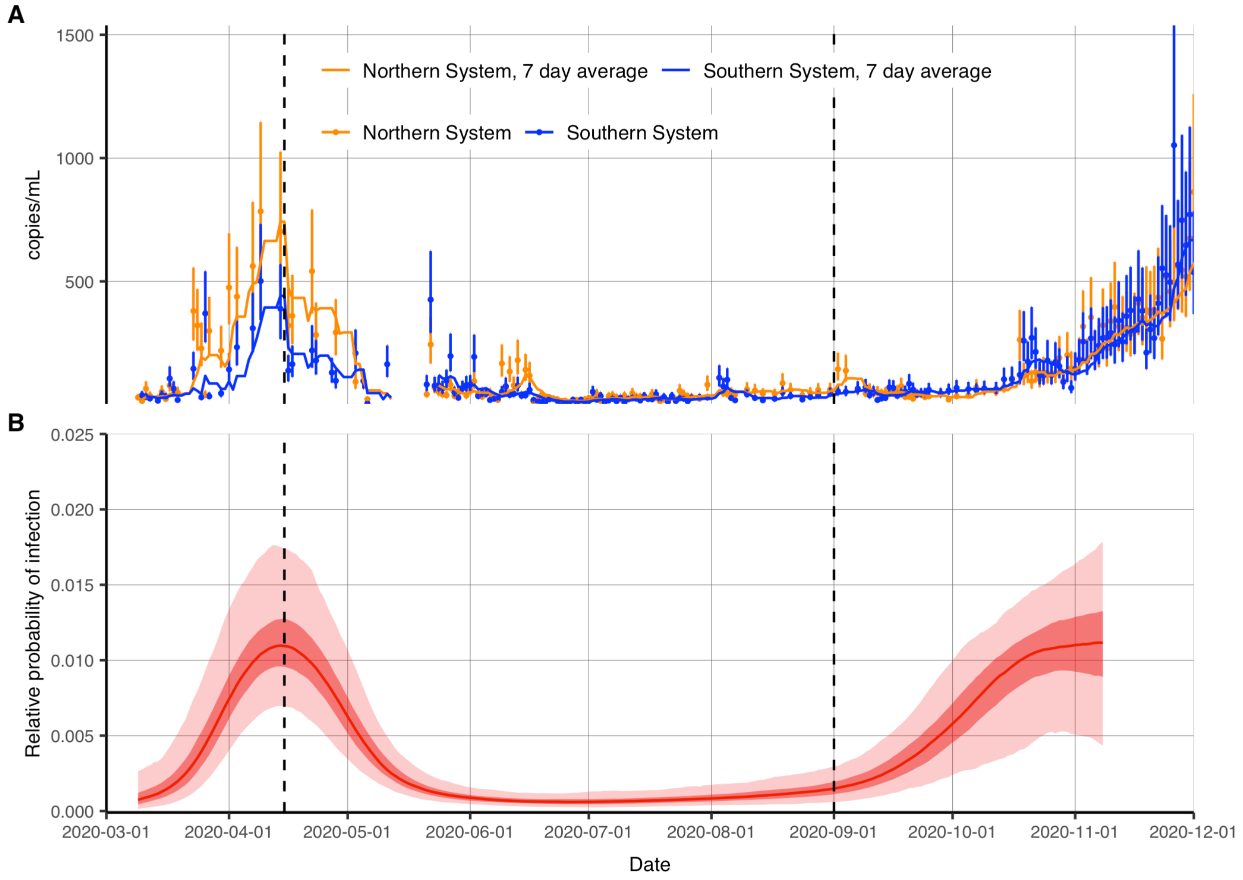


**Fig. S13. Epidemic trajectory inferred using Ct distributions from routine testing in Brigham & Women's Hospital in Massachusetts tracks viral loads in Massachusetts wastewater samples.** (**A**) SARS-CoV-2 viral RNA signal in Massachusetts wastewater over time (*33*). (**B**) As in Fig. 4, posterior distribution of relative probability of infection by date from a Gaussian Process (GP) model fit to all observed Ct values (ribbons show 95% and 50% credible intervals, line shows posterior median). Note that this is the same model fit, but the time axis extended back to 2020-03-01.

| Parameter | Description | Prior point estimate | Prior |
|---|---|---|---|
| **Viral kinetics and Ct value model** | | | |
| $t_{eclipse}$ | Time from infection to initial viral growth | 0.00 days | Fixed |
| $C_{zero}$ | Ct value at time of infection | 40.0 | Fixed |
| $t_{peak}$ | Time from initial viral growth to peak viral load | 5.00 days | Fixed |
| $C_{peak}$ | Modal Ct value at peak viral load | 20.6 (MNH) or 19.7 (BWH) | Normal(20.6, 2.00) or Normal(19.7, 2.00) |
| $t_{switch}$ | Time from peak viral load to secondary waning phase | 9.38 (MNH) or 13.3 days (BWH) | Normal(9.38, 3.00) or Normal(13.3, 3.00) |
| $C_{switch}$ | Modal Ct value at $a = t_{eclipse} + t_{peak} + t_{switch}$ | 33.0 (MNH) or 38.0 (BWH) | Normal(33.0, 1.00) or Fixed |
| $t_{LOD}$ | Time from infection until modal Ct value is equal to the limit of detection | Inf (plateau) | Fixed |
| $p_{addl}$ | Daily probability of detectability loss after $t_{switch}$ | 0.110 (MNH) or 0.103 (BWH) | Beta(11.9, 95.9) or Beta(10.5, 91.2) |
| $VL_{LOD}$ | Limit of detection of viral load ($\log_{10}$ RNA copies / mL) | 3.00 | Fixed |
| $C_{LOD}$ | Limit of detection of Ct value | 40.0 | Fixed |
| $\sigma_{obs}$ | Initial scale parameter for the Gumbel distribution until $a=t_{eclipse}+t_{peak}+t_{switch}$ | 5.15 (MNH) or 5.00 (BWH) | Normal(5.15, 0.50) or Normal(5.00, 0.50) |
| $s_{mod}$ | Multiplicative factor applied to scale parameter for the Gumbel distribution starting at $a = t_{eclipse}+t_{peak}+t_{switch}+t_{scale}$ | 0.400 (MNH) or 0.789 (BWH) | Fixed |
| $t_{mod}$ | Time from secondary waning phase until Gumbel distribution reaches its minimum scale parameter | 14.0 days | Fixed |

| SEEIRR and SEIR models | | | |
|---|---|---|---|
| $R_0$ | Basic reproductive number | Estimated (4 locations) | Log-normal(log(2.00), 0.60) and bound between (1.00, 10.0) |
| $t_0$ | Effective seed time | Estimated (4 locations) | Uniform(2020-03-01, 2020-05-11) |
| $I_0$ | Proportion infected at seed time | 0.002 (1 in 500) | Fixed |
| $1/\sigma'$ | Pre-detectable latent period | 2.00 | Log-normal(log(2.00), 0.30) |
| $1/a$ | Pre-infectious incubation period | 2.00 | Log-normal(log(2.00), 0.30) |
| $1/\gamma'$ | Infectious period (SEEIRR) | 4.00 | Log-normal(log(4.00), 0.60) |
| $1/w$ | Post-infectious detectable period (SEEIRR) | 11.0 | Log-normal(log(11.0), 0.30) |
| $1/\sigma$ | Incubation period (SEIR) | 4.00 | Log-normal(log(4.00), 0.25) |
| $1/\gamma$ | Infectious period (SEIR) | 4.00 | Log-normal(log(4.00), 0.50) |
| **GP model** | | | |
| $\nu$ | Maximum covariance between two time points | 1.50 | Fixed |
| $\rho$ | Rate of decline of the covariance between two time points as distance increases | 0.03 | Fixed |

**Table S1. Parameters used the viral load and cycle threshold (Ct) value distribution, the susceptible-exposed-infectious-recovered (SEIR) transmission model, the SEEIRR transmission model, and the Gaussian process (GP) model.** Note that different priors for the viral kinetics parameters are assumed for the Massachusetts Nursing Home (MNH) or Brigham & Women's Hospital (BWH) data.

**Movie S1. Multiple cross-sections of cycle threshold (Ct) values can be combined to improve the estimation of the epidemic trajectory over time.** Animation of epidemic trajectory estimation (bottom) using the Gaussian process GP model fit repeatedly to additional samples of observed cycle threshold (Ct) value data (top) in an ongoing simulated epidemic. The red line indicates the true daily per capita incidence of the simulated data.
